# Low-cost, local production of a safe and effective disinfectant for resource-constrained communities

**DOI:** 10.1101/2023.07.07.23292341

**Authors:** Andrea Naranjo-Soledad, Logan Smesrud, Siva RS Bandaru, Dana Hernandez, Meire Mehare, Sara Mahmoud, Vijay Matange, Bakul Rao, N Chandana, Paige Balcom, David Olugbenga Omole, Cesar Alvarez-Mejia, Varinia Lopez-Ramirez, Ashok Gadgil

**Affiliations:** Department of Civil and Environmental Engineering, University of California, Berkeley, Berkeley, California, United States of America; The Molecular Foundry, Lawrence Berkeley National Laboratory, Berkeley, California, United States of America; Department of Chemical and Nuclear Engineering, University of California, Berkeley, Berkeley, California, United States of America; VINYAS Architects, Urban Designers, Landscape Architects, Delhi, India; Centre for Technology Alternatives for Rural Areas, Indian Institute of Technology Bombay, Mumbai, Maharashtra, India; Centre for Emerging Technologies for Sustainable Development, Indian Institute of Technology Jodhpur, Jodhpur, Rajasthan, India; Engineering R&D, Takataka Plastics, Gulu, Uganda; Biosystems Engineering, Gulu University, Gulu, Uganda; Department of Civil Engineering, Covenant University, Ota, Ogun State, Nigeria; Department of Environmental Engineering, Tecnológico Nacional de México, ITS de Abasolo, Abasolo, Guanajuato, Mexico; Department of Biochemical Engineering, Tecnológico Nacional de México/ITS de Irapuato, Irapuato, Guanajuato, Mexico

## Abstract

Improved sanitation and hygiene depend on the accessibility and availability of effective disinfectant solutions. These disinfectant solutions are unavailable to many communities worldwide due to resource limitations, among other constraints. Safe and effective chlorine-based disinfectants can be produced via simple electrolysis of salt water, providing a low-cost and reliable option for on-site, local production of disinfectant solutions to improve sanitation and hygiene. We report on a system (herein called “Electro-Clean”) that can produce concentrated solutions of hypochlorous acid (HOCl) using only low-cost and now widely accessible materials. Using only table salt, water, graphite welding rods, and a DC power supply, HOCl solutions (∼1.5 liters) of 0.1% free chlorine (i.e. 1000 ppm) can be safely produced in less than two hours at low potential (5 V DC) and modest current (∼5 A). Rigorous testing of free chlorine production and durability of the Electro-Clean system components, described here, have been verified to work in multiple locations around the world by our project team, including microbiological tests conducted in two different countries to confirm the biocidal efficacy of the Electro-Clean solution as a surface disinfectant. We provide cost estimates for making HOCl locally with this method in the USA, India, and Mexico. Our findings show that Electro-Clean is an affordable alternative to off-the-shelf commercial chlorinator systems in terms of first costs (or capital costs), and cost-competitive relative to the unit cost of the disinfectant produced. By minimizing dependence on supply chains and allowing for local production, the Electro-Clean production process has the potential for improving public health by addressing the need for high-strength disinfectant solutions in resource-constrained communities.

## Introduction

Chlorine-based disinfectants are widely used in a variety of settings, including hospitals, public spaces, food preparation, and drinking water disinfection (1–3). The use of disinfectants dramatically reduces the risk of disease transmission and has many positive public health benefits, yet more than 25% of the world’s population does not have access to basic sanitation (4). Even when disinfectants are available, due to cost and limited supply chains, disinfectant solutions are often diluted excessively as a way to ration the solution, in turn making it a less effective disinfectant (5,6). On-site chlorine production and its utilization locally circumvents risks associated with chlorine storage and transport in bulk quantities (7). Electrochlorination is an electrochemical technique that enables the on-site production of chlorine-based disinfectants such as sodium hypochlorite and hypochlorous acid with ordinary table salt (sodium chloride) and electricity as inputs, thus avoiding supply chain issues (8). However, most commercially available electrochlorinators use expensive electrode materials, such as dimensionally stable mixed metal oxide (MMO) electrocatalyst anodes consisting of iridium and ruthenium oxides such as RuO2^-^ and IrO2^-^ (9). Limited global reserves of Ir and Ru and an increase in demand for ^M^MO electrodes in the Chlor-Alkali industry and other novel electrochemical applications ar^e^ responsible for the high prices of these metals ($100,000 per Kg of Ir and $20,000 per kg of Ru), resulting in limited availability, even in industrialized countries (10). While MMO anodes have high efficiency in electrochemical chlorine generation, they make electro-chlorinators costly and inaccessible to resource-limited communities. Additionally, electrochlorination studies and efforts have primarily focused on applications to drinking water disinfection and thus the production of dilute concentrations of free chlorine that are insufficient for surface-disinfection applications (11,12).

Prior efforts to produce chlorine-based disinfectant solutions using low-cost materials include a student team from MIT that focused on the use of carbon rods obtained from carbon-zinc batteries, a simple saline solution, and a recycled plastic cup as the container (13). Although this design was quite inexpensive and effective at producing free chlorine, it required the extraction of carbon rods from zinc-carbon batteries. In our experience with collaborators in various countries in non-academic settings, we found significant resistance and concern by lay people to breaking open a carbon-zinc battery to extract the carbon rod, despite reassurances of its harmlessness. This user feedback highlighted the importance of minimal complexity, one of the key characteristics of adoption rate as defined by Rogers and other researchers on the Diffusion of Innovations (14). We abandoned further improving the design developed by the team at MIT since its complexity would significantly lower the rate of adoption. To overcome potential resistance, we put our focus on a reactor design that would minimize complexity to the end-user, even if that slightly increased the cost of production.

In this paper, we report on a simple electrochemical process (herein called “Electro-Clean” for short) that strives to eliminate these economic and accessibility barriers, improving access to best practices of hygiene and sanitation in resource-poor environments. Electro-Clean has low capital and operating costs, and produces a powerful, safe-to-use hypochlorous acid disinfecting solution that has been approved for such use by WHO, EPA, and CDC (15–17). The Electro- Clean process can produce HOCl solutions at concentrations sufficient for addressing surface disinfection in high-contamination settings such as hospitals, producing 1.5 liters solution with 0.1% free chlorine (i.e. 1000 ppm) in less than two hours, operating at low potential (5V DC) and modest current (around 5A). The development and testing of Electro-Clean involved collaboration with our co-authors and other project partners in seven countries spanning three continents, exemplifying how this process can be replicated in a variety of contexts with locally available materials.

### Electrolysis for chlorine generation

Chlorine-based disinfectants such as sodium hypochlorite or hypochlorous acid can be produced via the electrolysis of sodium chloride (NaCl) solutions (herein referred to as “salt water”) (18). The Electro-Clean production system, schematically shown in Fig 1, uses graphite welding rods, salt water, and a switched mode power supply (SMPS) to produce a high-concentration of chlorine-based disinfectant that can be pH adjusted for maximum biocidal activity.

**Fig 1.**
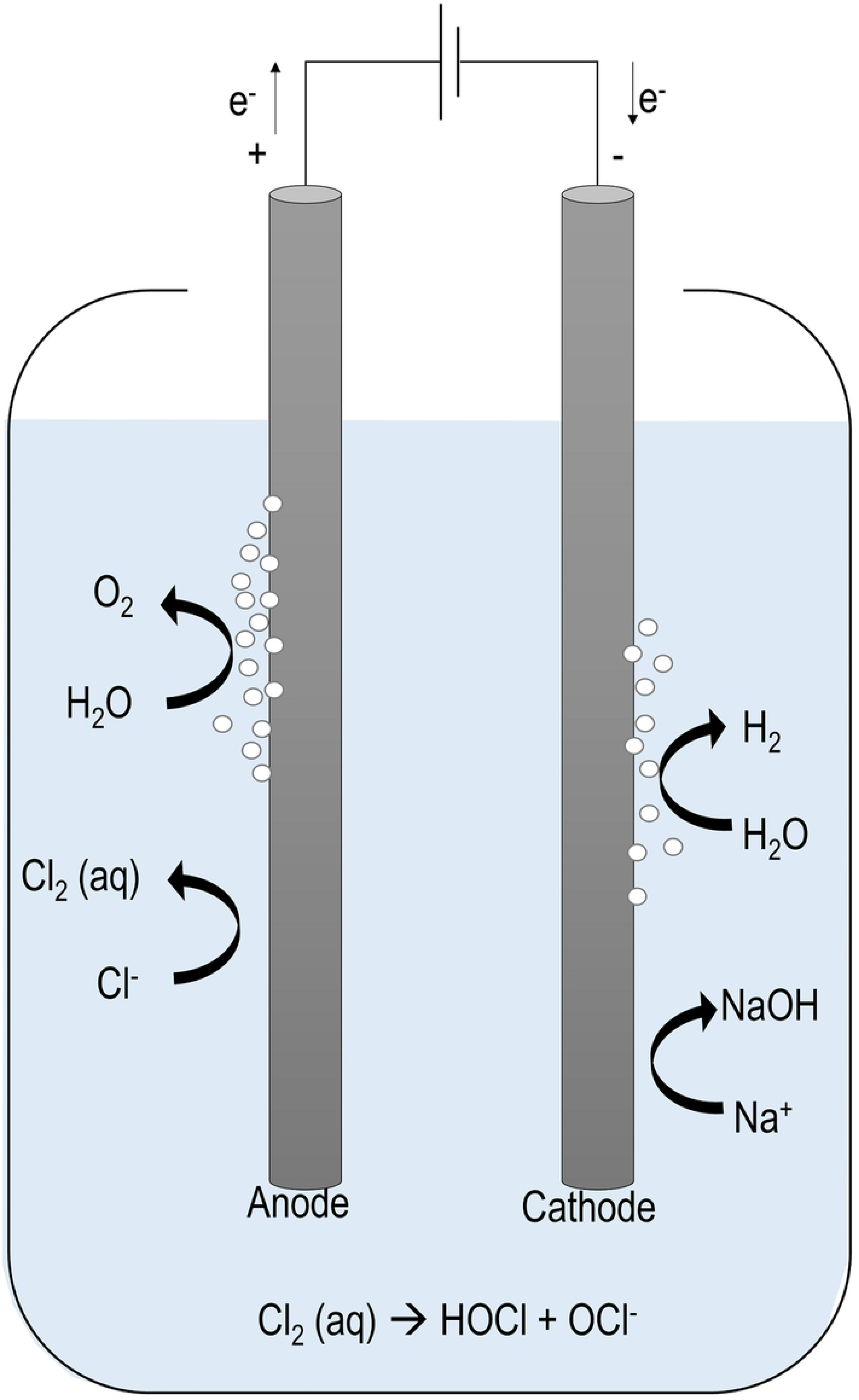
Schematic of Electro-Clean reactor system. The Electro-Clean reactor system consists of two carbon gouging rods as electrodes, 30,000 mg/L sodium chloride dissolved in tap water as the electrolyte solution, and an externally applied potential that drives the reaction to produce chlorine gas on the anode, which gets immediately hydrolyzed to produce hypochlorous acid and/or hypochlorite in the electrolyte.

Design considerations and material selection for creating an effective disinfectant at an affordable price involved making trade-offs between efficiency of free-chlorine production, electrode durability, and cost. Lower cost and more readily available materials that we recommend are associated with moderately reduced chlorine production compared to more high-end electrodes and have shorter lifetimes than costly electrode materials. However, even with these disadvantages, the HOCl produced with this process is highly effective and both capital cost and total cost remain affordable compared to its alternatives. Other design considerations included optimizing parameters that lead to higher electrical currents (i.e. Amperes delivered) to the reactor, without increasing the cost too significantly, and are summarized in Section S5. High current can be achieved with high salinity and/or reducing interelectrode distance. A high current is desirable as it leads to shorter electrolysis times. Operating the reactor at a high current also comes with trade-offs as it leads to a more rapid degradation of the electrodes. High salinity on the other hand requires high salt content and dramatically increases the cost of production. These trade-offs were considered in our extensive preliminary testing (not reported here) leading to the selection of materials and configurations that we recommend and report on for reactor design.

The carbon graphite welding rods, as shown in Fig 1, allow for chlorine production at the anode with continuous subtle degradation due to oxidation of the electrode material over time (19–21). Although disintegration of the electrode is undesirable, using a low-cost, widely available consumable electrode material such as carbon is still preferred. Other common electrode materials, such as stainless steel, are undesirable as they are not stable in the presence of the chloride ion and will rust when a current is applied (22–24). Therefore, stainless steel electrodes can only be used as the cathode, limiting the operational flexibility and ability to reverse polarity to prolong the longevity of the electrodes. Other high-performing electrodes, such as Iridium- Ruthenium Mixed Metal Oxides (MMO), are not only quite expensive, but also locally unavailable in most parts of the world, and thus not considered viable for the Electro-Clean system.

Overall, the Electro-Clean process is simple, requiring only affordable materials and minimal operator knowledge. This allows for on-site generation as opposed to dependency on supply chains, market availability, and financial capacity to meet high up-front costs. The hypochlorous acid solution, once generated using the Electro-Clean process, slowly returns to salt and water over a period of months. The rate of returning back to a salt solution is higher at elevated storage temperatures, exposure to strong UV light, and higher HOCl concentrations (25–27). Due to this instability, the use of the solution for disinfection is recommended within a week or so from its production date. This ensures its high disinfectant potency and limits the conversion of the disinfectant back to a salt solution.

### pH dependence of chlorine-based disinfectants

The efficacy of chlorine-based disinfectants is pH dependent. At a pH below 4, chlorine exists primarily as dissolved chlorine gas. As shown in Fig 2, at a pH of 6, chlorine exists in water mostly as hypochlorous acid (HOCl), and as pH increases above 8, the dominant species shifts to hypochlorite (OCl^-^) ions. These different species have varying disinfecting powers, with HOCl being 80 to 100 times more effective at bacterial inactivation than the hypochlorite ion (OCl^-^) (28). The amplification of biocidal properties in HOCl compared to NaOCl illustrates the importance of pH adjustment for chlorine-based disinfectants (29).

**Fig 2.**
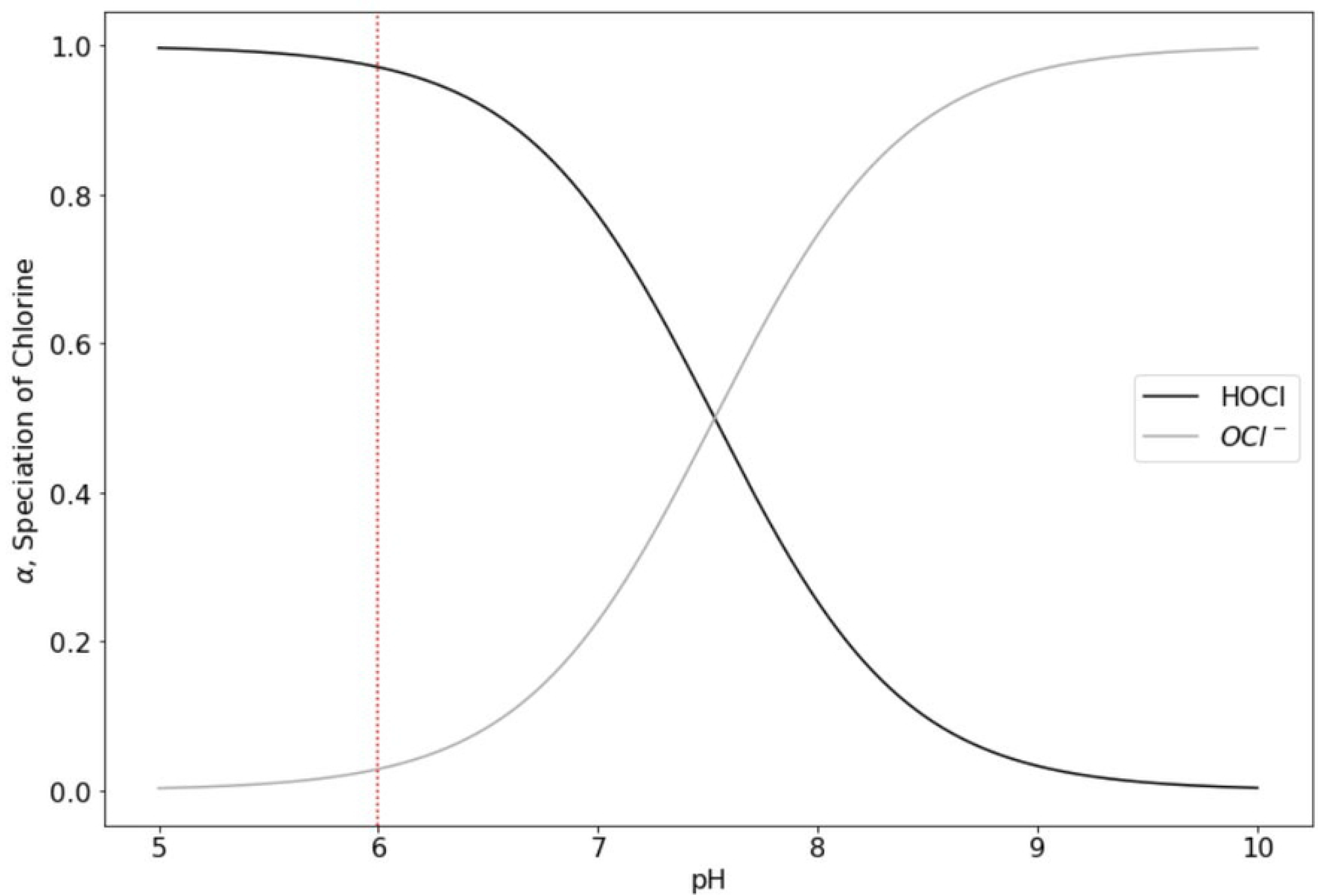
Speciation of aqueous chlorine species (represented by the fractional alpha value) as a function of pH, around circumneutral pH values. In the pH range shown above, the predominant species is hypochlorous acid at the lower end and shifts to the hypochlorite ion at the upper end. The vertical line shown at pH 6 is the target pH value for the disinfectant solution produced by the Electro-Clean process, where hypochlorous acid is the predominant species. This figure was produced using published chlorine speciation equations (30). Alpha represents the fractional concentration of a chlorine species over the total chlorine concentration.

Chlorine gas hydrolyzes to hypochlorous acid quite rapidly, and proceeds as follows:

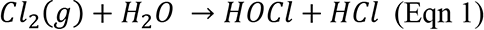

Hypochlorous acid can proceed to dissociate to the hypochlorite ion, as follows:

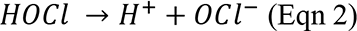

Graphite electrodes were historically used as anodes in the Chlor-Alkali industry for chlorine generation. After the development of MMO anodes, graphite electrodes were discontinued in the Chlor-Alkali industry because of their high overpotential and gradual anodic consumption by parasitic oxygen evolution reactions at the industry-relevant current densities (>100 mA/cm^2^) (31–34). In this work, we employed graphite anodes for on-site chlorine generation over MMO anodes because of their low cost and widespread availability. Furthermore, low operating current densities (<10 mA/cm^2^) and low cell voltage (<6 V) are chosen in the Electro-Clean process to preferentially promote Cl2 released at the anode.

### Multi-use of produced hypochlorous acid solution

#### Surface disinfection

Disinfecting high-touch surfaces is common practice in many public spaces. Upon the arrival of the COVID-19 pandemic in early 2020, this cleaning measure became an important way to reduce the risk of fomite transmission of the virus. Preventing nosocomial infections acquired in healthcare settings is another relevant application for chlorine-based disinfectants. Especially in low-resource settings where supply chains may be unreliable and disinfectants in short supply, on-site generation of chlorine-based disinfectants has been shown to dramatically improve essential hygiene practices (35). These practices include regular disinfection of surfaces, regular hand washing and sanitizing, and always having high-strength disinfectants readily available for the prevention of hospital-acquired infections which are particularly relevant to safe births for women and their newborns (35,36).

#### Topical applications

The high-strength disinfectant produced by the Electro-Clean process can be diluted to appropriate lower concentrations and then used as a non-alcohol-based hand sanitizer. Numerous studies have explored the use of dilute (0.01%, or 100 ppm) solutions of hypochlorous acid as an antiseptic for wound healing, and in dental and eye-care settings, among other clinical applications (37–39).

#### Drinking water disinfection

Chlorine has been used to disinfect drinking water for many decades, and current drinking water standards stress the importance of pH as it relates to its bactericidal and virucidal properties (40,41). An important parameter for calculating disinfectant dosage for the chlorination of drinking water is the “CT” value, typically expressed in units of mg-min/L, where “CT” is the product of the residual disinfectant concentration “C”, and the contact time “T”, in the water being disinfected (42). For the context of this work, when added to contaminated, non-turbid source water in an amount equal to 1/250 (v/v) of the source water volume, and held for 4 hours, the high-strength (1000 ppm) hypochlorous acid solution could be used to disinfect drinking water from some bacterial and viral contaminants. This dosing would yield a CT value of 960 mg-min/L. Assuming temperatures are no less than 10 degrees Celsius, source water pH is in the range of 6 to 9, and at least half of the free chlorine dosed remains present as residual, then this CT value of 960 mg-min/L would be sufficient for 3-log inactivation of Giardia cysts, which are some of the most resistant to chlorination (43).

All the above uses of HOCl could be accessible to resource-limited communities via the Electro- Clean production process for low-cost, on-site generation of the chlorine-based disinfectant.

Instructions for how to produce Electro-Clean are open-access and available online and have been translated into at least four languages (44). Public seminars and global communities of practice also exist on the topic of decentralized chlorine production (45).

## Materials and methods

After several months of iteration and experimentation with different materials and designs, we arrived at the materials and methods described here. All sub-optimal designs are omitted for brevity. Larger, more complex designs can be found in Section S5.

### Reactor materials and designs

#### Electrolyte

The electrolyte solutions were prepared using either reagent-grade NaCl, or commercial-grade NaCl, and local municipal tap water. To mimic real-world conditions, electrolytic chlorine generation with solutions prepared using commercially available iodized (Morton iodized salt in the USA) and non-iodized (Diamond Crystal granulated plain salt in the USA) salts were compared, and the resulting chlorine production showed no significant difference. To ensure easy reproducibility of our results by others, non-iodized salt was used in all experiments reported here.

Local tap water was used for all experiments. In the Berkeley area, tap water is supplied by the East Bay Municipal Utility District, and its composition is cited in their water quality report (46). An electrolyte solution of 30,000 mg/L NaCl (a similar salinity to that of seawater) was used for all experiments unless stated otherwise.

#### Power supply

With the advent of computers, the internet, and LED lighting, Switched-Mode Power Supplies (SMPS), have become ubiquitous. They perform high-efficiency conversion of grid power into a 5-Volt supply with high current capacity (e.g. 20 A or 40 A). With mass production, their prices have rapidly come down. We chose SMPS supplies over dry-cell batteries and full wave rectifiers because the SMPS are lower cost, higher efficiency, higher reliability, and also have higher safety compared to a home-built full wave rectifier. A Switched-Mode Power Supply (SMPS) was used to supply the low potential required to drive the electrolysis reaction. The SMPS (MEISHILE brand, Amazon.com) supplied 5 Volts DC, with a maximum output current of 40 Amperes.

#### Electrodes

Although MMO electrodes produce chlorine with nearly 100% Faradaic efficiency, they are unavailable and highly expensive in the context of our intended users. We chose consumable graphite gouging electrodes due to their low cost and widespread availability, even though their Faradaic efficiency is significantly lower, at about 20%.

We selected uncoated bare carbon gouging rods to serve as both the anode and cathode. To remove the loose carbon particles on the electrode surface, they were rinsed with tap water and brushed with a nylon brush before use. The carbon gouging electrodes had a diameter of 9.52 mm (3/8 in) and a length of 30.5 cm (12 in) (McMaster-Carr).

In some countries, like Mexico and Nigeria, carbon gouging rods were only found with copper coating. Therefore, chlorine production using copper-coated carbon gouging rods was also investigated in this study. We found that the copper coating could be simply peeled off with small pliers (Fig 3), leaving copper coating only at the tip of the rod where the electrical connections are made. A small section of the stripped copper coating could also be repurposed for use at other electrical connection points given that the smooth, conductive material facilitates a strong electrical connection and thus a stable current delivery.

**Fig 3.**
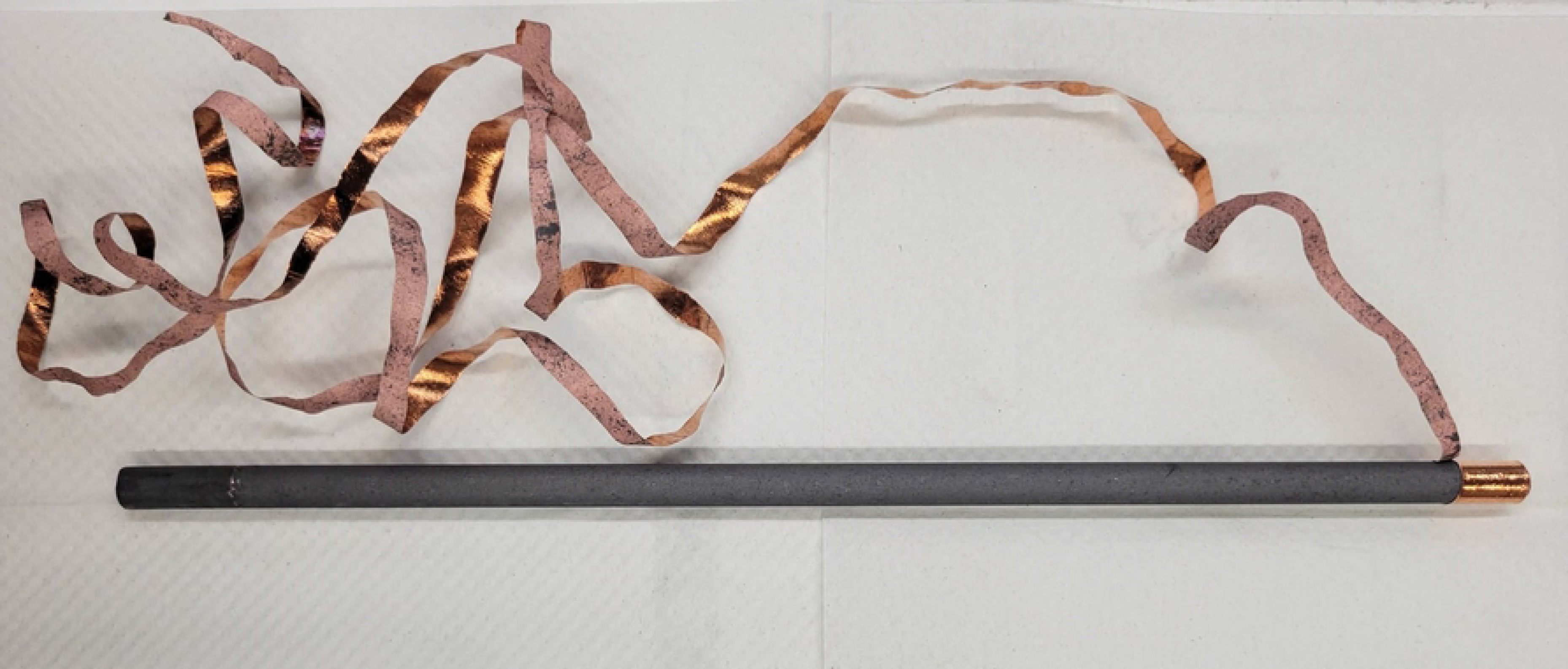
**Copper peeled off as a winding strip from the copper-coated carbon gouging rod.**

#### Filtration of carbon slurry

During electrolysis, electrodes gradually disintegrate and particles from the disintegrating carbon gouging rod form a visible slurry. Multiple filter materials were tested to remove these suspended carbon particles from the final hypochlorous acid solution. The materials tested were nylon cloth, geotextile, and coffee filters. For the nylon cloth and geotextile, the filter material was secured around the anode during electrolysis to prevent the carbon particles from escaping into the bulk solution. For the coffee filter, the solution at the end of electrolysis was poured over the filter for particle removal. The concentration of free chlorine was measured with and without the use of filters for comparison.

#### Reactor assembly and operation

The simplest reactor assembly for the Electro-Clean process consisted of a 1.5-liter soda bottle, carbon gouging electrodes, a multimeter, an SMPS, and a 30,000 mg/L table salt electrolyte solution prepared from tap water. A 12-gauge power cord with a three-prong plug on one side was used to connect the SMPS to the power outlet. The multimeter was connected in series to measure current, and a 12 AWG insulated electrical wire was used for making connections between the electrodes and the SMPS. The Electro-Clean reactor assembly is shown in Fig 4A. The two carbon gouging electrodes were aligned in parallel and held tightly together with rubber bands; and rubber bands were also used as spacers between the electrodes to prevent short-circuiting, as shown in Fig 4B.

**Fig 4.**
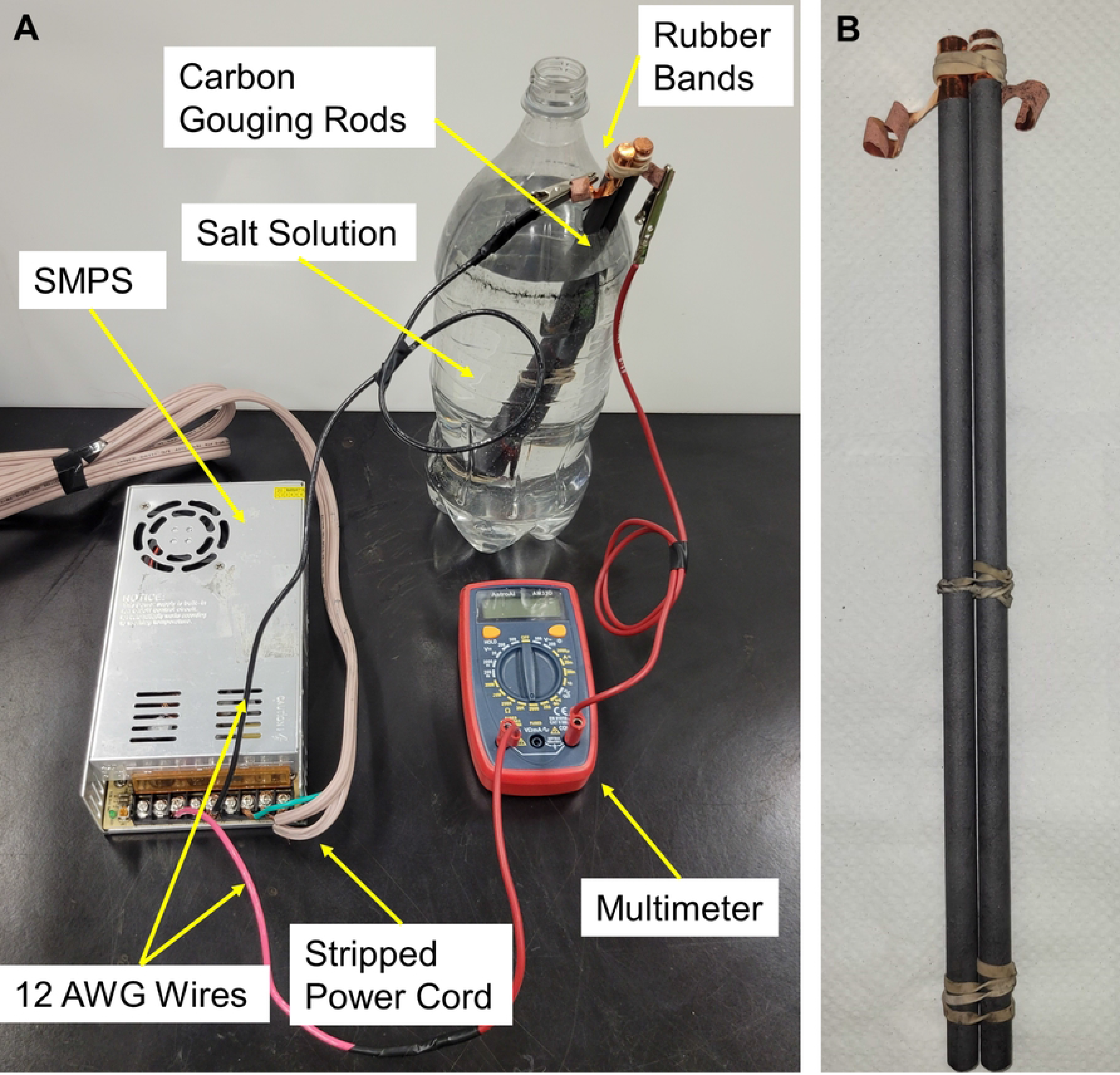
Digital picture of the Electro-Clean assembly. (A) Electro-Clean batch reactor assembly, and (B) Carbon gouging electrode assembly.

Batch experiments reported in this section used two 30.5 cm (12 inches) long uncoated carbon rods with 1 cm (3/8 inch) diameter; 25.4 cm (10 inches) of their length submerged into a 1.5-liter bottle with 1.35 L of 30,000 mg/L NaCl solution. As seen in Fig 4A, we cut an opening near the top part of the bottle’s sidewall to insert the carbon electrode rods. The reactor was operated for 90 minutes with an externally supplied direct current of 5 Volts. This resulted in a supplied current of about 5 Amps, depending on the internal resistance of the circuit connections.

#### Other size scales considered for Electro-Clean

In addition to the 1.5-liter soda bottle design, four other designs were explored: 1) a smaller, household-scale design producing 250 mL of concentrated HOCl solution, 2) a larger, community-scale design capable of producing 15 L of dilute HOCl solution, 3) a co-axial electrodes assembly, and 4) a reactor using a 1.5-liter soda bottle, but with multiple, parallel electrode assemblies, capable of producing the concentrated HOCl in a shorter time. For both the 250 mL and 15 L designs, stainless steel, and carbon rods were studied as the cathode and anode, respectively. A more detailed description of these designs is in Section S5.

### Quantitative measurements

#### Chlorine concentration measurements

The UV-Vis spectrophotometer (Hach DR6000) was used to measure the free chlorine concentration in the generated disinfectant through colorimetry at a wavelength of 530 nm. The N,N-diethyl-p-phenylenediamine, or DPD, reagent sachet (Hach brand) was added to 10 mL of the diluted disinfectant solution, producing a pink color when free chlorine is present. The intensity of the pink color, accurately measured with the spectrophotometer, led to the quantitative accurate measurement of free chlorine in the solution.

#### pH adjustment

Various materials were tested for their suitability to adjust the pH of the electrolyte and/or disinfectant solution, including distilled white vinegar (Heinz, 5% acidity), lemon juice, and lime juice. Early experiments showed that although the natural acidity of the lemon and lime juice could decrease the pH of the disinfectant solution to the desired value of 6, they were unsuitable due to their high content of organic matter which leads to the disappearance of free chlorine (47). In these preliminary experiments where lemon or lime juice were used to lower the pH, complete disappearance of free chlorine was observed in as little as five minutes, reducing the disinfectant solution to only a simple salt solution with a lemon essence, and thus further investigation with lemon and lime juice was not pursued nor recommended. This rapid consumption of free chlorine by lemon and lime solutions has been well documented by others (12).

For all experiments presented herein, unless stated otherwise, distilled white vinegar (i.e. dilute acetic acid) was used to adjust the pH. Acetic acid was selected for these experiments due to its wide market availability. Hydrochloric acid (HCl, also known as muriatic acid), is a less expensive and equally effective alternative for lowering pH when distilled white vinegar is unavailable. However, in many communities, hydrochloric acid is a widely regulated chemical due to safety concerns. We acknowledge that muriatic acid may have yielded slightly different observations and conclusions than ours with acetic acid in regards to pre and post pH adjustment, due to the possible interaction of acetic acid with electrolysis products. We also acknowledge that vinegar is likely more expensive than dilute HCl. Nevertheless, we accept these limitations given that vinegar is more widely available and therefore more suitable for the intended Electro- Clean users.

#### Faradaic efficiency tests

Faradaic efficiency of an electrode is calculated as the ratio between the observed free-chlorine production (measured with the UV-Vis spectrophotometer) and the theoretical free chlorine production (calculated using Faraday’s Law). This parameter provides unique insight into the efficiency of chlorine production by the Electro-Clean process. A thorough explanation of the Faradaic efficiency calculations is presented in Section S1.

#### Durability of electrodes over time

During the electrolysis of a brine solution, carbon anode electrodes undergo oxidation, slow mechanical disintegration, and eventual failure (19,20). This process occurs not only on the surface of the electrodes but also within their high-porosity structure. In order to slow the disintegration of the carbon rods during electrolysis, a technique of filling the pores with light mineral oil (commercial name Johnson & Johnson Baby Oil) was explored. Each carbon gouging electrode was submerged in light mineral oil for 20 hours prior to use. Only the tips of the electrodes, where the connections to the power supply were made, were left unsubmerged.

The two performance parameters used to evaluate the durability of the electrode rod were 1) free chlorine concentration, and 2) current. The 1.5-liter reactor system design was used to observe the durability of the bare carbon rods as well as mineral-oil-soaked carbon rods. The electrolyte was a 30,000 mg/L NaCl solution. The electrolyte volume in the reactor was 1.35 L. A peristaltic pump was used to continuously feed the reactor with electrolyte, and excess electrolyte was continuously removed from the reactor. The flow rate of 0.25 mL/s led to a hydraulic residence time of 90 minutes for the electrolyte in the reactor volume. All samples were taken at the outlet. See Fig S10 for more detail on the continuous-flow assembly.

#### Effect of electrolyte pH on free chlorine production

The effect of electrolyte pH pre-electrolysis on free chlorine generation was studied. We compare an electrolyte solution with a semi-neutral pH of 8 to one with a slightly acidic pH of 5. Distilled white vinegar was used to adjust the pH of the slightly acidic electrolyte. For both conditions (electrolysis at pH 5, and electrolysis at pH 8), the post-electrolysis pH was measured and adjusted to 6 using distilled white vinegar to ensure that all available chlorine was present as HOCl. The 1.35-liter Electro-Clean process assembly in batch mode was used for this study. The electrolysis time was 90 minutes and the electrolyte was 30,000 mg/L NaCl.

#### Free chlorine decay over time

Prior literature has noted that HOCl solutions are best stored in a dark, cool place, and in glass or plastic (not metal) containers to slow down the loss of free chlorine over time due to reconversion back to a salt solution (25–27). We studied free chlorine decay over time of the produced disinfectant solution when stored at room temperature in a dark closet. We also considered the material of the storage container (glass versus plastic), and the presence of vinegar (used for pH adjustment).

The disinfectant solution was produced following the Electro-Clean process in a 2-liter plastic soda bottle using tap water (local to Berkeley). The free chlorine concentration of the produced disinfectant was 915 ± 35 ppm as Cl2. This was then diluted in half (∼440 ppm), which is the concentration at which the chlorine decay experiments began.

The diluted HOCl solution (440 ppm, 500 mL in each container) was stored in two glass bottles and two plastic bottles. About 3 mL of vinegar was added to one plastic bottle and one glass bottle. This lowered the pH from 8.8 to 6.4 ± 0.1 in the plastic bottle and glass bottle. All four bottles were closed to prevent evaporative loss and periodically monitored for free chlorine over a month duration (34 days).

#### Reproducibility in other communities worldwide

To ensure that the Electro-Clean process performed similarly in other contexts worldwide, co-authors in India, Mexico, Nigeria, and Uganda replicated the Electro-Clean process using reasonable substitution of locally available materials. While the main components of the Electro- Clean process remained the same (i.e., gouging-carbon electrodes, 5-Volt/20-Amp SMPS, tap water, salt, vinegar, and a plastic container), the manufacturer of each component varied from community to community. In the case of Nigeria, the co-authors used dilute HCl for pH adjustment instead of white vinegar. Table S2 summarizes the manufacturer specifications of the materials used by co-authors outside the USA.

#### Microbiological assays to evaluate the biocidal effect of the produced hypochlorous acid solution

To validate the disinfecting capabilities of the Electro-Clean solution, microbiological assays were conducted for its application on various high-touch material surfaces in public spaces and laboratory settings in Mexico, and laboratory settings in India. In each case, a hypochlorous acid solution was prepared by the Electro-Clean process, as described in Section 2.1.5. It was then diluted to the concentration of intended use (in this case, of approximately 250 ppm for surface disinfection), and subsequently adjusted with distilled white vinegar to pH 6. The materials and methods used for the microbiological studies performed in Mexico and India can be found in Sections S7 and S8.

#### Comparative cost analysis

To quantify the cost of the Electro-Clean disinfectant production system, and to understand which items contribute to that total cost, an analysis evaluated both the capital and operating costs of Electro-Clean. To conduct this economic assessment and comparative analysis, costs were obtained from three countries (USA, India, Mexico) for all reactor components. The Electro-Clean system was also compared against commercially available electro-chlorinators and locally available chlorine-based disinfectant products.

## Results and discussion

### Chlorine production by carbon electrodes in batch processes

In a single anode/cathode configuration, repeated and averaged over triplicates, the following values were observed: 3.82 ± 0.14 Ampere average current, 17.1 ± 0.3 percent Faradaic efficiency for free chlorine production, and 962 ± 20 ppm of total free chlorine production over 90-minute electrolysis times in 1.35 L batch volumes.

### Repeated batch experiments for insight on electrode durability

Prior to conducting the long-term durability experiments on the electrodes, the batch process was repeated to gain insight into electrode lifespan and facilitate experiential planning for the long-term experiments. These preliminary experiments were set up in a 1.75 L electrolyte volume made from tap water and 30,000 mg/L dissolved salt. The power supply was an SMPS providing a constant 5 Volts. The wires from the SMPS were wrapped around a piece of the peeled-off copper coating that was still attached to the welding rod. The initial pH was 8.71 ± 0.06. The average concentration of free chlorine after 10 batches (each of 30 min electrolysis time) was 459 ± 38 ppm as Cl2. The turbidity was the highest in the first experiment at 9.73 NTU. After the first batch, the turbidity was 1.35 ± 0.29 NTU, with an average current of 4.36 ± 0.16 Amperes. Results for Carbon-Copper welding rods are presented in Fig S8. These results were obtained prior to those presented in Section 3.2.

### Durability of electrodes over time

Three long-term experiments were carried out in the laboratory to quantify the longevity of the bare carbon gouging electrodes, and to study the effect of soaking the carbon electrodes in mineral oil to minimize disintegration. The results showed Faradaic efficiency remained relatively constant, even right before the anode ruptured. Current, on the other hand, gradually decreased over time as the anode degraded. A reduction in current and visual thinning of the anode (due to disintegration) were indicators that the anode was soon to rupture.

The data as a function of charge passed (Coulombs) for the bare and mineral oil-soaked electrodes from all three trials can be found in Section S6. The average values for the three trials for bare and mineral-oil-soaked carbon electrodes are shown in Fig 5. Although the mineral-oil-soaked electrodes seemed to operate at a slightly higher average current as seen in Fig 5A, the difference in current is within the margin of error. Soaking the carbon rods in mineral oil allowed for a higher amount of charge to be sustained, resulting in a longer useful lifetime for the carbon electrode. Fig 5C demonstrates that the mineral-oil-soaked carbon rods can last approximately 20 hours (about 45%) longer than the bare carbon rods when operated at the same current. However, soaking the rods in mineral oil lowered the free chlorine concentration by 31%, as seen in Fig 5D. Correspondingly, soaking the rods in the mineral oil also lowered the Faradaic efficiency.

**Fig 5.**
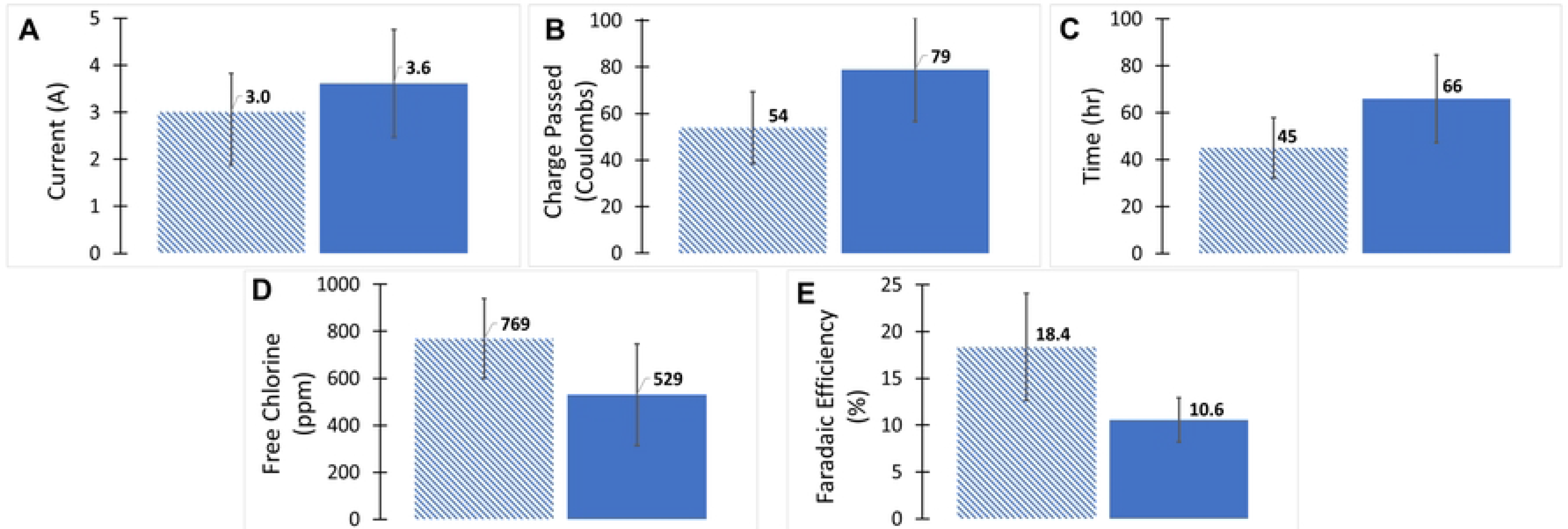
Long-term (∼50 hours) performance of 1.5-liter Electro-Clean set-up using bare carbon electrodes (dashed column) and mineral-oil-soaked carbon electrodes (solid column). (A) Average current during long-term operation, (B) Average charge passed in Coulombs on the electrodes before failure, (C) Average time in hours of the anode lifespan, (D) Average free chlorine concentration produced for a residence time of 90 minutes in a 1.35-liter volume, (E) Average Faradaic efficiency for producing free chlorine throughout the long-term operation.

**Fig 6.**
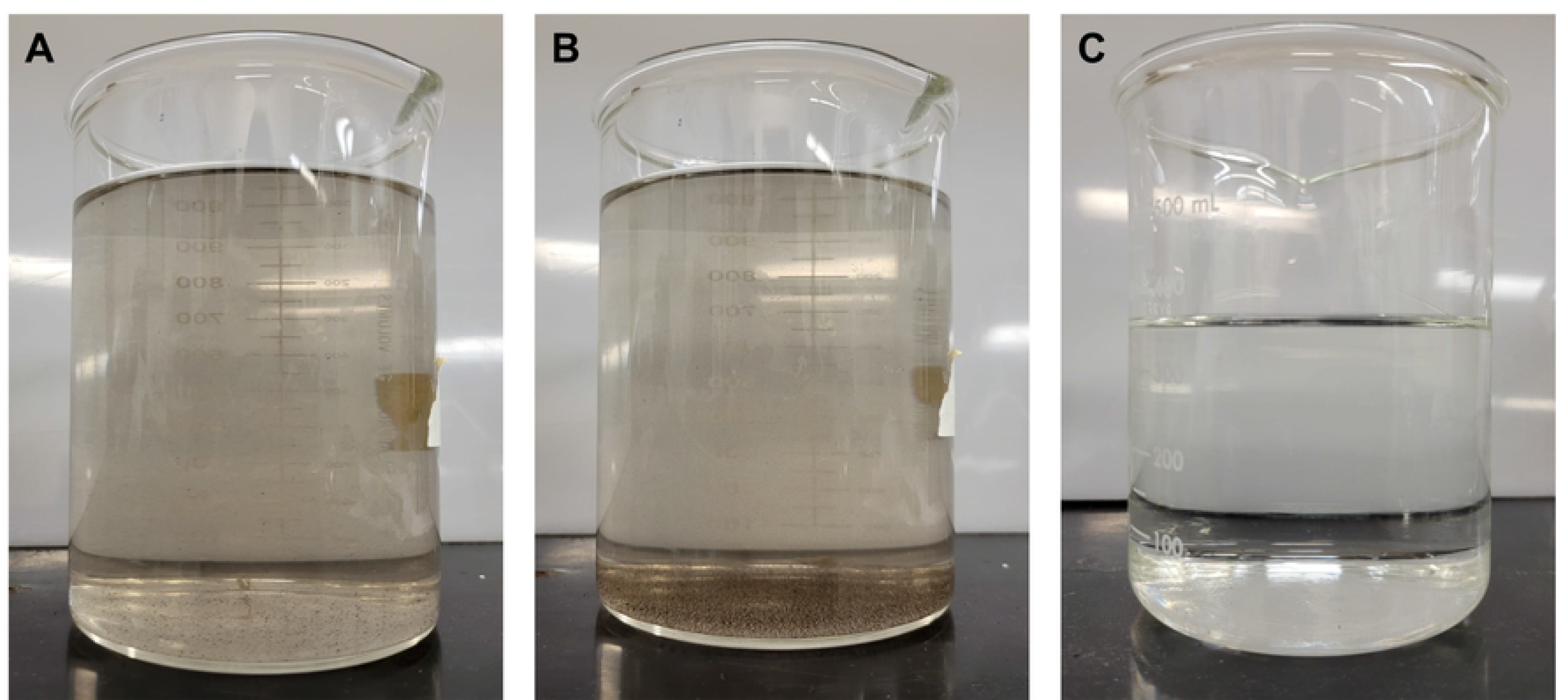
Digital images of Electro-Clean solutions after electrolysis. (A) Freshly generated HOCl solution, (B) produced HOCl solution after 1 hour of settling, and (C) produced HOCl solution after filtering through two coffee paper filters.

This could be due to the mineral oil promoting other side reactions, such as oxygen evolution, at the anode.

Findings from the long-term experiments described here and presented in Fig 5 suggest that although the electrode life can be prolonged by submerging the anode in mineral oil, the advantage is offset by a reduction in production efficiency of free chlorine. As a result, we do not recommend this approach. However, future studies could investigate other low-cost, readily available materials to enhance the durability of carbon gouging electrodes, as it is an important aspect of improving the sustainability and cost-effectiveness of this disinfectant production process.

### Electrode disintegration during electrolysis

As noted earlier, the electrolysis reaction causes anodic carbon rod disintegration, which releases carbon particles in the electrolyte solution (20,48–50). In this process, the amount of free chlorine produced was not affected, yet the cloudy physical appearance was undesirable for a cleaning product.

We found that a fine-mesh nylon cloth wrapped around the anode carbon rod does not hold the carbon particles within the cloth for the allotted reaction time. A fine-mesh geotextile wrapped around the carbon rod during the reaction successfully prevented the carbon particles from dispersing into the solution, however, the free chlorine yield was greatly reduced. Filtering the electrolyte post-electrolysis through two layers of ordinary coffee filter paper effectively separated the slurry particles from the electrolyte, and this filtering did not significantly affect the concentration of free chlorine. After passage through the double coffee filters, the visibly cloudy solution became visibly clear and the chlorine concentration slightly decreased from its initial value of 450 ± 77 ppm as Cl2 to a final value of 438 ± 50 ppm.

### Effect of electrolyte pH on free chlorine production

As shown in Table 1, regardless of the initial solution pH, at the end of each electrolysis, the final solution pH increased to the same value of approximately 8.4 for both the neutral and slightly acidic initial electrolysis conditions. As a result, the total amount of acetic acid required for pH adjustment for a final solution with pH 6 remained about the same between the two conditions, in the range of 25 to 28 mL of acetic acid for 1.35 L electrolyzed or to-be electrolyzed salt water, as noted in Table 1. This observation is likely due to the buffering capacity of NaOH that is created during the 90-minute electrolysis. Relative to the standard deviations, a modest (20%) increase is observed in chlorine production under slightly acidic conditions relative to neutral conditions, as seen in Table 2. Vinegar usage was relatively unchanged between the two set-ups, and chlorine production did not differ significantly.

**Table 1.** White distilled vinegar usage for pH adjustments before and after electrolysis in acidic and neutral conditions.

|  | **Electrolysis at a near-neutral pH (8.4)** | **Electrolysis at a slightly acidic pH (5.0)** |
| --- | --- | --- |
| **pH before electrolysis** | 8.4 (no adjustment) | 5.0 (adjusted with acetic acid) |
| **pH after electrolysis** | $8.5 \pm 0.2$ | $8.3 \pm 0.1$ |
| **pH of final HOCl solution** | $5.8 \pm 0.2$ (adjusted with acetic acid) | $5.8 \pm 0.2$ (adjusted with acetic acid) |
| **Total volume of vinegar added (mL)** | $28 \pm 1.2$ | $25 \pm 4.7$ |

**Table 2.** Chlorine production in neutral and slightly acidic electrolysis conditions.

|  | **Average Current**<br>**(Amp)** | **Faradaic Efficiency for**<br>**Free Cl<sub>2</sub> (%)** | **Total Cl<sub>2</sub> (ppm)**<br>**immediately after**<br>**electrolysis (pH ~8.4)** |
| --- | --- | --- | --- |
| **Electrolysis at a**<br>**near-neutral pH**<br>**(8.0)** | 4.87 ± 0.76 | 18% ± 1.9 | 1297 ± 113 |
| **Electrolysis at a**<br>**slightly acidic**<br>**pH (5.0)** | 4.46 ± 0.85 | 15% ± 1.0 | 982 ± 177 |

Our results suggest that regardless of a neutral or slightly acidic solution pre-electrolysis, pH adjustment after electrolysis may be crucial to ensuring a final pH of ∼6 in chlorine solution disinfectants. This step is missed by many commercially available electro-chlorinators that direct users to adjust the pH only before electrolysis.

With the aim for Electro-Clean to be an easy, user-friendly process, it is recommended to conduct the electrolysis at neutral pH and make pH adjustments after electrolysis to minimize the number of pH adjustment steps required. It is also recommended to make pH adjustments closer to the time of use to minimize chlorine decay over time as HOCl is less stable than NaOCl.

### Free chlorine decay over time

For the non-acidified storage in both plastic and glass containers, the loss of free chlorine was less than 5% after 34 days, and the pH stayed about the same at 8.7 and 8.8, respectively. In the case of vinegar-acidified storage, the loss of free chlorine was 20% in the plastic bottle, and 16% in the glass bottle after 34 days. This is consistent with the prior literature stating that HOCl (present at an acidic pH value) is more prone to decay than the OCl^-^ ion (present at a higher pH value). At the end of the 34 days, the pH levels were 5.7 for the plastic bottle and 5.9 for the glass bottle.

As shown in Fig 7, over the duration of one month, the decay of the free chlorine concentration never exceeded 20% in the plastic or glass containers stored in a dark space. However, to maximize the disinfectant strength and maintain freshness, it is recommended that either the hypochlorous acid solution be stored in a dark, cool storage room, or that the solution’s pH be lowered only upon use.

**Fig 7.**
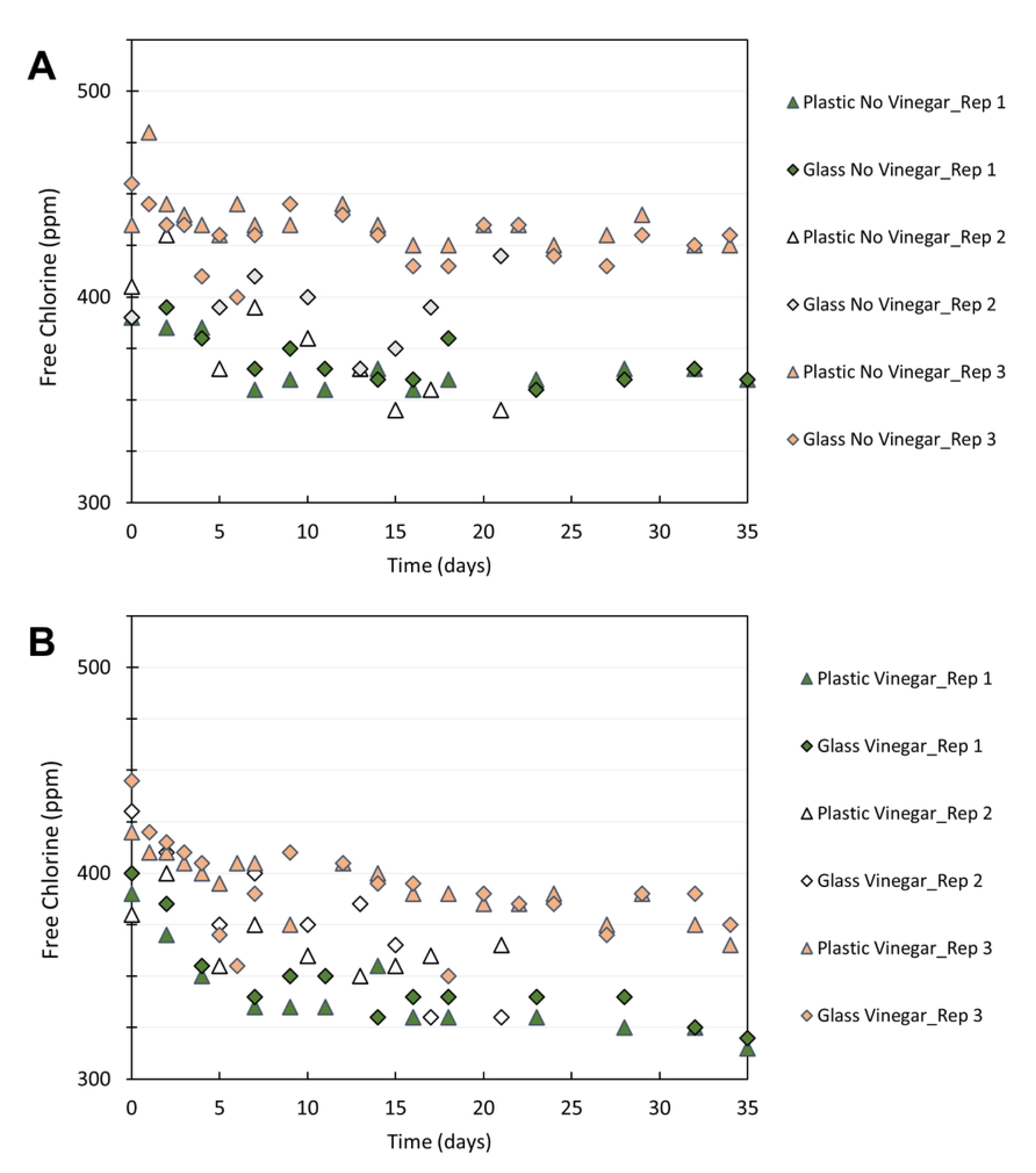
Free chlorine decay over time for stored Electro-Clean solutions. (A) Free chlorine in Electro-Clean solution without acidification, at approximately pH 8.7 stored in glass and plastic bottles, and (B) Free chlorine in Electro-Clean solution with vinegar-acidification, at approximately pH 6.4 stored in glass and plastic bottles.

### Microbial assays

The results of the microbial assays can be found in Sections S7 and S8, along with a detailed discussion of the findings. The results in Table S4 and Figure S19 suggest that the ∼250 ppm hypochlorous acid solution had similar disinfecting power to bleach 1% (v/v) and ethanol 70% (v/v) on various material surfaces.

Although the literature suggests that 300 ppm of HOCl is an adequate concentration for surface disinfection, due to the limited quality assurance and control of a DIY product outside controlled, laboratory conditions, like the Electro-Clean process performed outside a laboratory, it is recommended to use a higher concentration of the HOCl solution to perform disinfection comparably to ethanol 70% and bleach 1%.

### Implications of reactor design and reproducibility of the Electro- Clean process

The design of the Electro-Clean process was an iterative process that involved careful consideration of technical and social barriers that could affect the diffusion of the technology in various regions, communities, and contexts. It also included substantial user feedback during the design process. This critical step of technology design and innovation was done in partnership with our co-authors in communities and at universities across the globe.

Throughout these iterations, a handful of other alternative reactor components were considered. Although the final design for the Electro-Clean reactor uses an SMPS for the power supply, other options such as power supplies for LED lights or simple phone chargers were also considered.

These alternatives were ultimately rejected due to lower market availability, or due to the low current they could deliver (i.e. less than 1 Ampere), which led to a very low chlorine production rate. Extraction of the graphite core in dry-cell batteries was also initially considered for use as electrodes. However, user feedback quickly led us to conclude that people were uncomfortable with the safety of cutting open dry-cell batteries.

Alternative electrode materials were also considered in response to feedback on the difficulty of procuring certain electrode materials in some countries. Ultimately, graphite electrodes, also known as carbon gouging rods, were selected for use in the Electro-Clean process. Carbon gouging rods are widely used, even in small workshops (e.g. for automobile body repair shops), wherever steel welding is practiced. They were found to be low-cost and widely available for our project team members in India and Mexico, and could also be ordered via web-based suppliers like Amazon. Stainless steel plates were considered for use as the cathode, and most applicable for scaled-up applications of the Electro-Clean system. More details on these scaled-up and alternative reactor configurations are presented in Section S5.

Our co-authors and some of their collaborators within their countries, in both academic and non-academic settings, reproduced the Electro-Clean process using materials from local manufacturers and with local tap water, noting only small differences from those tested at the laboratory at UC Berkeley.

These slight differences in materials, as outlined in Table S2, used by our project team produced hypochlorous acid solutions at concentrations high enough for surface disinfection (i.e., 300 ppm as HOCl), if not higher. Table 3 summarizes the free chlorine concentrations achieved by our co-authors with their DIY Electro-Clean reactors before dilution and pH adjustment.

**Table 3.** Results of production of hypochlorous acid solution using Electro-Clean by co-authors in India, Mexico, Nigeria, and Uganda.

|  | Soda Bottle Scale (1.5-liter) |  |  | Community Scale (15-liter) |  |  |  |
| --- | --- | --- | --- | --- | --- | --- | --- |
|  | India<sup>(a)</sup> | Mexico | U.S.A. | India<sup>(b)</sup> | Nigeria | Uganda | U.S.A. |
| **Electrolysis time (min)** | 94 | 30 | 90 | 240 | 240 | 180 | 90 |
| **Volume of Electrolyte (L)** | 1.5 | 1.3 | 1.35 | 15 | 15 | 15 | 15 |
| **Average** | 9.42<sup>(c)</sup> | 1.2 | 3.02 | 15.8 | 4.82 | 7.5 | 6.65 |

| **current (A)** |  |  |  |  |  |  |  |
| --- | --- | --- | --- | --- | --- | --- | --- |
| **Average Free Chlorine Concentration (ppm)** | 1896<sup>(d)</sup> | 486 | 769 | 500 | 514 | 640 | 259 |
| **Volume of vinegar to lower pH ~6.0 (mL)** | 90 | 5 | 30 | 75 | 5<sup>(e)</sup> | 110 | 75 |

<sup>a</sup>Results from co-author Vijay Matange.
<sup>b</sup>Results from co-authors in IIT Bombay.
<sup>c</sup>Higher current was likely achieved due to larger diameter (1.5-cm) rods. See Table S2 for details.
<sup>d</sup>Result estimated assuming 18% Faradaic efficiency. Free chlorine measuring kit not available.
<sup>e</sup>Hydrochloric acid (HCl) was used for pH adjustment.

It should also be noted that the availability of chemicals for pH adjustment varied among co-authors and other project partners. Dilute muriatic (i.e., hydrochloric) acid, was an attractive choice for lowering pH without introducing other functional groups that could potentially interfere with chlorine production. However, this chemical proved difficult to obtain in non-academic settings due to safety concerns and was thus ruled out for recommended use in the Electro-Clean process. Vinegar is a very weak acid that is marketed as a food product and thus widely available in most countries and contexts. Vinegar does introduce the carboxylic acid group, but no effects on the disinfectant solution’s potency were observed.

### Cost analysis of disinfectant production with Electro-Clean

Table 4 and 5 show the detailed capital and operating costs of the 1.5-liter scale Electro-Clean system. As expected, a significant fraction (more than 50%) of capital costs for Electro-Clean are attributed to the SMPS. A higher amperage (5 V, 20 A) power supply costs were used in the cost estimates; however, a lower amperage power supply can decrease the capital costs significantly. In the USA, operating costs to yield the Electro-Clean solution was $0.15 per liter at 800 ppm free chlorine concentration. The 800 ppm is representative of an average free chlorine concentration produced during the long-term experiments presented in Fig 5. Corresponding operating costs in India and Mexico are $0.09 per liter and $0.11 per liter, respectively. In India and Mexico, consumable costs of carbon rods contributed significantly (more than 50%), whereas in the USA, the operating costs are dominated by the costs of salt.

**Table 4.**
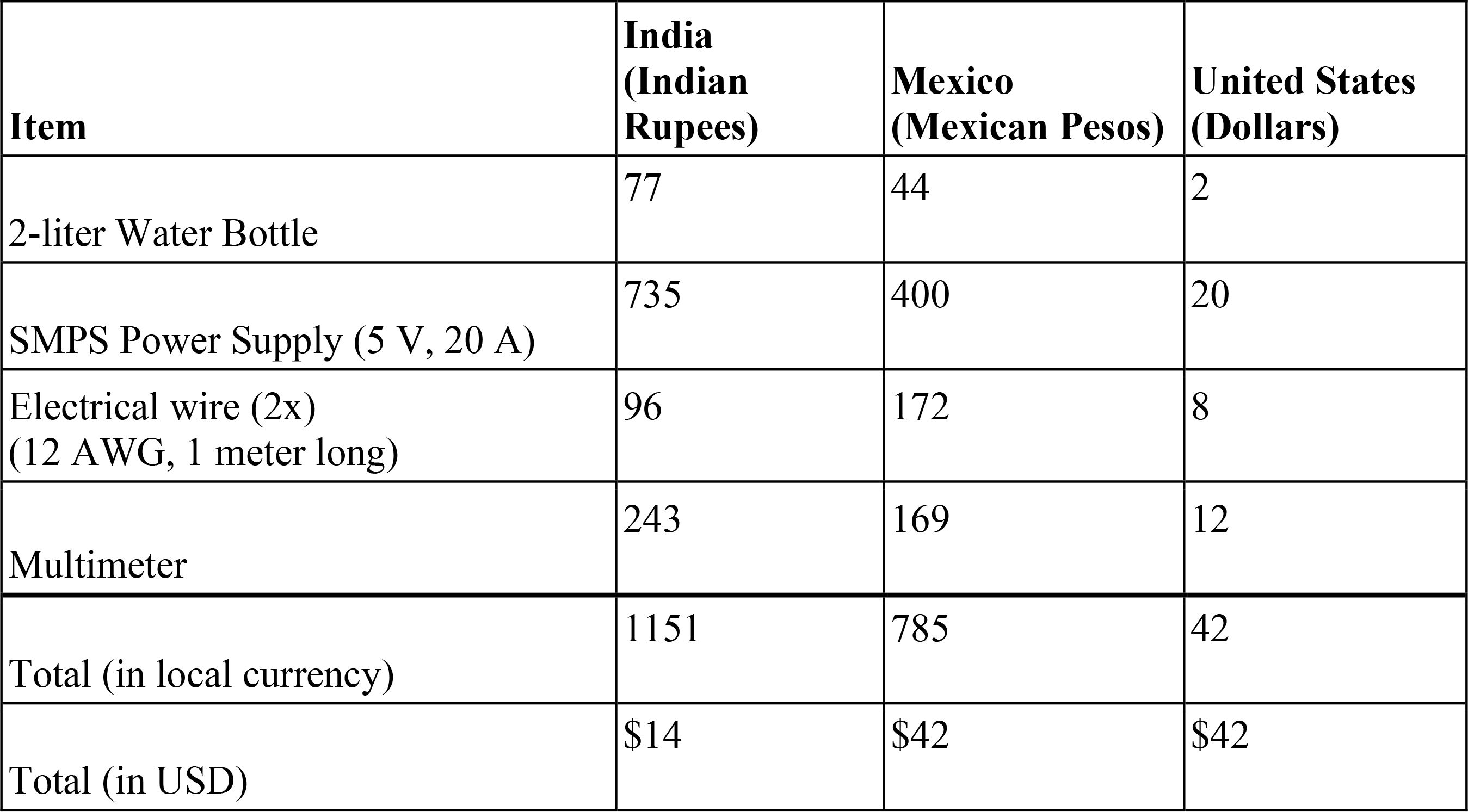
Capital Costs of 1.5-liter Scale Electro-Clean System.

A small variability in the capital and operating costs of Electro-Clean between India, Mexico, and the USA is attributed to the relative price differences in the various components as shown in Tables 5 and 6, respectively. With a 1.5-liter scale Electro-Clean system, nearly 60 liters of Electro-Clean solution at 800 ppm of free chlorine can be produced before the pair of carbon rods (with periodic polarity reversal) are completely exhausted. Electro-Clean is an affordable technology, with its capital costs being among the lowest for existing electro-chlorinator technologies in the market for personal, household, and large-scale applications. Table 6 shows the comparative cost analysis of Electro-Clean with other electro-chlorinator technologies.

**Table 5.** Operating Costs of 1.5-liter Scale Electro-Clean System for producing 1.35 L solution of 800 ppm Free Chlorine.

| Item | India<br>(INR per L) | Mexico<br>(Pesos per L) | United States<br>(USD per L) |
| --- | --- | --- | --- |
| Carbon Welding rods (2x)<br>(~5 mm diameter, 30 cm long) | 4.93 | 1.15 | 0.02 |
| Vinegar | 0.75 | 0.65 | 0.10 |
| Sodium Chloride | 1.69 | 0.31 | 0.03 |
| Electricity | 0.12 | 0.03 | 0.00 |
| Total (in local currency per L) | 7.48 | 2.14 | 0.15 |
| Total (in USD per L) | \$0.09 | \$0.11 | \$0.15 |

**Table 6.**
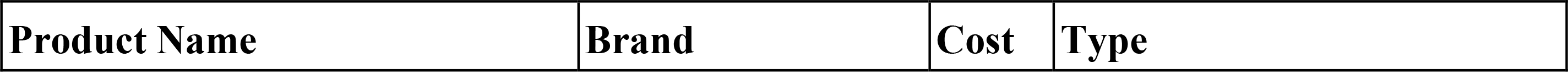
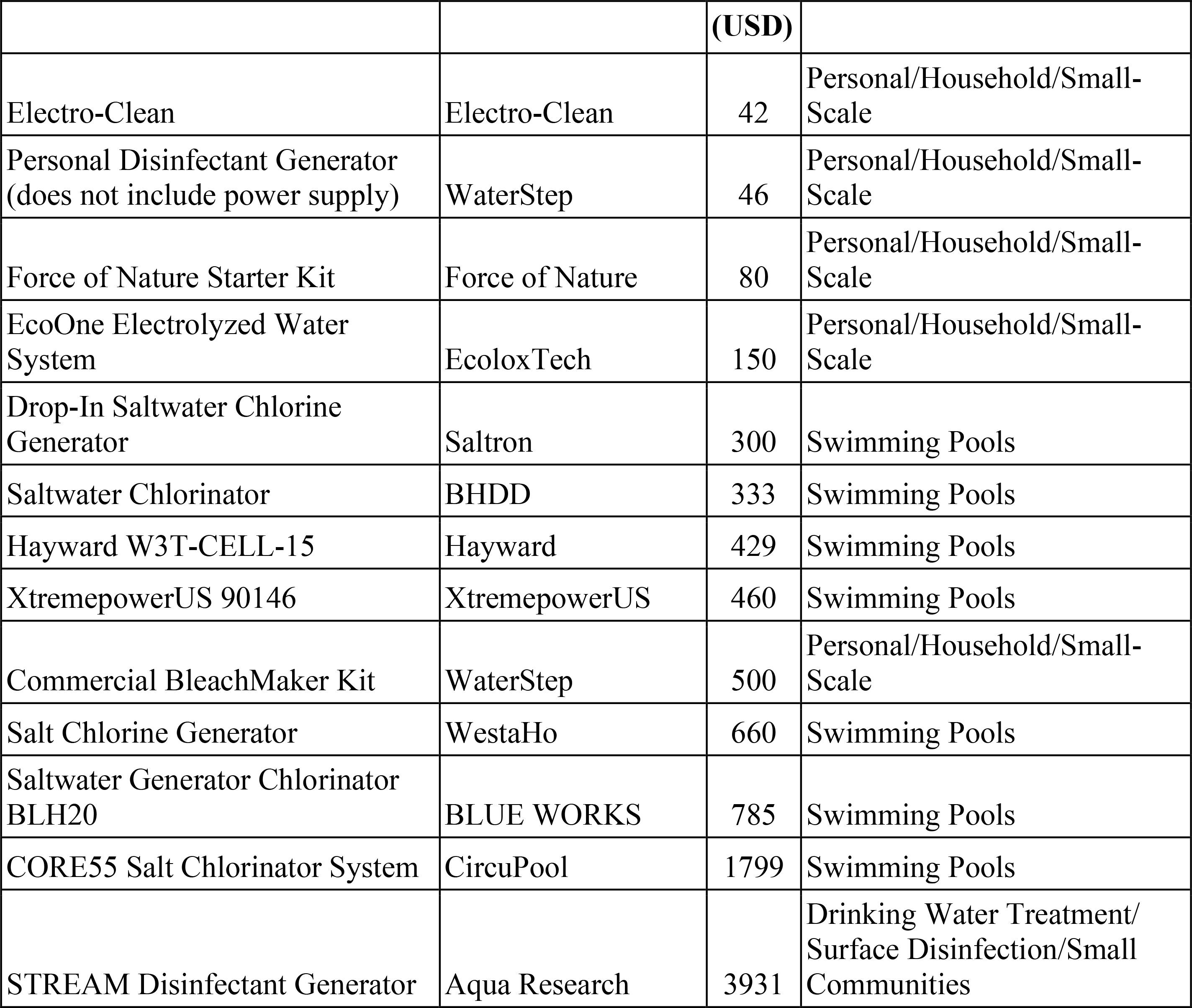
Capital cost comparison of available existing electro-chlorinators. Prices as of April 2023.

| Product Name | Brand | Cost | Type |
| --- | --- | --- | --- |

|  |  | (USD) |  |
| --- | --- | --- | --- |
| Electro-Clean | Electro-Clean | 42 | Personal/Household/Small-Scale |
| Personal Disinfectant Generator (does not include power supply) | WaterStep | 46 | Personal/Household/Small-Scale |
| Force of Nature Starter Kit | Force of Nature | 80 | Personal/Household/Small-Scale |
| EcoOne Electrolyzed Water System | EcoloxTech | 150 | Personal/Household/Small-Scale |
| Drop-In Saltwater Chlorine Generator | Saltron | 300 | Swimming Pools |
| Saltwater Chlorinator | BHDD | 333 | Swimming Pools |
| Hayward W3T-CELL-15 | Hayward | 429 | Swimming Pools |
| XtremepowerUS 90146 | XtremepowerUS | 460 | Swimming Pools |
| Commercial BleachMaker Kit | WaterStep | 500 | Personal/Household/Small-Scale |
| Salt Chlorine Generator | WestaHo | 660 | Swimming Pools |
| Saltwater Generator Chlorinator BLH20 | BLUE WORKS | 785 | Swimming Pools |
| CORE55 Salt Chlorinator System | CircuPool | 1799 | Swimming Pools |
| STREAM Disinfectant Generator | Aqua Research | 3931 | Drinking Water Treatment/<br>Surface Disinfection/Small<br>Communities |

To reinforce the affordability of on-site chlorine generation compared to the purchase of ready-to-use commercial bleach products, we compare the cost of the Electro-Clean solution against the cost per liter of the commercial bleach product. When comparing these costs, it is important to note that the disinfecting properties of HOCl (the active ingredient in Electro-Clean) is much stronger than that of NaOCl (the active ingredient in bleach), and thus greater volumes of the bleach solution are needed to meet the same levels of disinfection. When using chlorine-based disinfectants for surface disinfection, it is typically recommended to use 200 ppm as HOCl or 2000 ppm (0.2%) as NaOCl (1,51).

The standard price of bleach in the USA is approximately $1 USD per liter (Clorox, 7.5% as sodium hypochlorite, Amazon) (52). When diluting the bleach stock to 0.2% for use as a surface disinfectant, the cost becomes $0.03 per liter. This can be compared to $0.04 per liter of the diluted 200 ppm HOCl solution produced by Electro-Clean (i.e., a fourth of $0.15 per liter of 800 ppm, as shown in Table 5). In Mexico, the price of bleach is approximately $0.65 USD per liter (Clorox, 7.5% as sodium hypochlorite, Mercado Libre) (53). After dilution, the price lowers to $0.018 USD per liter. It is important to note that cost is not the only barrier to accessing disinfectants in low-resource or rural areas, and that vulnerable supply chains and transportation can play a role in accessibility. As such, comparisons in Tables 4 through 6 are between Electro- Clean and other electro-chlorinators, as opposed to the chlorine-based disinfectant itself (e.g. diluted bleach).

Commercially available electro-chlorinators were compared against Electro-Clean on the basis of capital cost. A comparison of operating and maintenance costs would be challenging due to most products not making that information publicly available to the public.

The low capital cost of the Electro-Clean system in comparison to other commercial alternatives, as shown in Table 6, increases accessibility to on-site chlorine generating devices.

### Understanding the context of intended application in communities

Collaborations were critical to the success of this project, as they allowed for rapid two-way exchange of knowledge, and allowed sharing of findings in a variety of contexts. We deployed the production and scale-up of Electro-Clean in more than five countries, in settings that included classrooms, households, and community organizations. Furthermore, collaboration among co-authors led to a collective awareness and understanding of the complex social and technical barriers associated with technology adoption of Electro-Clean. The availability of electrode materials, for example, varied greatly and presented challenges for selecting suitable substitutes where the recommended materials were unavailable.

Although low-cost disinfectants have always been in demand, the Electro-Clean project was initiated in response to the COVID-19 pandemic. The timeliness of these efforts is highlighted by the very rapid scale-up by some of our co-authors at their respective institutions. For example, at Covenant University in Ota, Nigeria, co-author Omole, and his students rapidly obtained permissions and administrative support for substantial scale-up efforts of Electro-Clean for practical use at the height of the COVID-19 pandemic in early 2020. Nigeria went in and out of lockdowns in 2020 as the different waves of the pandemic raged, and the demand for hand sanitizers and disinfectants soared. Thus, the advent of Electro-Clean as an affordable and safe disinfectant was a welcome contribution. The team at Covenant University responded to the sudden increased demand for surface disinfectants with a rapid scale-up of Electro-Clean, and also used the opportunity for community health promotion. They scaled up the Electro-Clean reactor design to a community scale, using 20-liter (5-gallon) plastic buckets, further described in Section S9, to meet the large quantities of disinfectant in demand.

Over a six-month period, the Covenant University team produced over 45 liters of the concentrated Electro-Clean disinfectant (i.e., >800 ppm as HOCl), which was diluted by adding three parts water (thus totaling 180 liters of ready-to-use disinfectant at 200 ppm as HOCl). The team distributed over five hundred 250-mL labeled spray bottles to the University faculty and students for use on high-touch surfaces in homes, classrooms, and offices. The labels contained brief user instructions, a statement of regulatory approval by the University authorities, and information on where the disinfectant solution could be refilled. The contents of every spray bottle were quality controlled, with a final step of measuring the free chlorine concentration prior to distribution. Photos of the scale-up effort at Covenant University can be found in Section S9.

User acceptance of the disinfectant was promoted with attractive labeling of the spray bottles. Use and popularity of the high-touch surface disinfectant noticeably declined as the COVID-19 pandemic waned, airborne transmission was identified as the overwhelmingly dominant route of transmission, and people mostly returned to business as usual.

Hurdles associated with regulatory approval by local governments and public perception were encountered by the co-author and team from Takataka Plastics in Gulu, Uganda, where the on- site generated hypochlorous acid was not officially recognized as an effective and approved disinfectant. Because the new disinfectant was not approved by the country’s National Bureau of Standards, some prospective users did not believe the new disinfectant was effective.

Furthermore, the local prospective users often did not trust the chlorine-based disinfectant because it did not smell like the alcohol-based disinfectants to which they were accustomed. This challenge emerged during the scale-up of the Electro-Clean production and distribution efforts since the Uganda National Bureau of Standards only has a category for alcohol-based disinfectants and does not have tests or procedures for evaluating non-alcohol-based disinfectants. To address these regulatory hurdles, that team’s future work involves developing a test protocol that could be proposed to the Bureau for testing of the chlorine-based disinfectant product for approval.

As noted earlier, COVID-19-related interest in Electro-Clean surged during the early months of the pandemic and declined as it became clear that aerosol transmission was the dominant mode of transmission. Nevertheless, the large unmet need among resource-poor communities and healthcare facilities for reliable access to surface disinfection was emphasized in the series of open virtual workshops organized starting in 2022 by PATH (45). Establishing partnerships with hospitals and health clinics operating in rural, low-resource settings could be a next step as the demand for low-cost disinfectants will always be relevant.

## Conclusions

The development of the Electro-Clean process was not a “top-down” or “isolated” approach, but instead a culmination of rapid design feedback from our partners (some of them co-authors of this paper), as we experimented with different materials and reactor sizes, landing on a design that made the most sense for a large fraction of our intended users. The selection of reactor components built off known knowledge of salt electrolysis for chlorine production, but prioritized criteria such as availability, cost, and simplicity over achieving maximum free chlorine production. The health benefits of a safe and effective disinfectant, even when produced with only a moderately efficient method (like Electro-Clean), vastly outweigh the costs when compared to the “do nothing” approach. Although the Electro-Clean system uses commonplace welding carbon rods for electrodes, instead of expensive state-of-the-art materials, substantial concentrations of free chlorine (more than 1000 ppm as Cl2) can be produced with this approach at affordable costs. Prior to pH adjustment, the Electro-Clean process makes bleach (i.e., sodium hypochlorite, a chlorine-based disinfectant) at about five times less efficiency (Faradaic efficiency of ∼20%) than off-the-shelf chlorine generator products, causing electricity costs to jump five-fold. However, electricity costs are a small fraction of the operating cost. So, a Faradaic efficiency of only ∼20% (translating into 5 times more electricity use) is a great tradeoff against much lower cost of electrodes (graphite versus Mixed-Metal-Oxide electrodes). Because the final product is so low-cost and can be locally made, Electro-Clean is a worthwhile and accessible alternative.

In terms of biocidal activity and disinfectant performance, the hypochlorous acid solution produced by the Electro-Clean process was observed to be effective in reducing colony-forming units of bacteria on contaminated high-touch surfaces. Its effectiveness was comparable to standard disinfectant solutions (e.g. diluted bleach, and 70% ethanol). The low-cost, widely available carbon gouging electrodes used in the Electro-Clean process were shown to be reasonably durable, with a lifetime of 45 operating hours per anode and the solution concentration remained relatively stable for up to a month when properly stored in the dark and at room temperature.

Even though the Electro-Clean process is robust, effective, and low-cost, there are still inevitable challenges and social barriers to technology adoption. Through collaboration with our co-authors and partners in communities around the world, we consistently observed that people are reluctant to adopt “complex” technologies - “the simpler the better”. We also observed that implementing this technology at a household level is quite challenging, while technology adoption at a community level where one person is in charge of producing the disinfectant for other users is more well-received. To lower the barrier of entry to comfortably interact with this technology, public access to instructional documents and videos on how to replicate this work were placed online as a small stride towards addressing the large-scale problem of limited availability and accessibility to disinfecting solutions (44).

By further developing this on-site HOCl production technology, using locally available materials, and bulking up outreach efforts, we hope that the Electro-Clean process can be a reliable solution for communities and organizations in need of a low-cost disinfectant to improve public health.

## Data Availability

All data necessary for reproduction, validation, reanalysis, new analysis, and reinterpretation are included either in the main manuscript or the supporting information.

## Acknowledgments

We sincerely thank the collaborating institutions who made this work possible, for allowing Electro-Clean to reach a larger audience and potentially empower communities around the world to produce commercial-strength disinfectants in low-resource settings. Those institutions include Covenant University in Nigeria, Higher Technological Institute of Abasolo Mexico, Higher Technological Institute of Irapuato Mexico, IIT Bombay, Jadavpur University, Potential Energy, Baobab Learning Center in Mali, Gulu University in Uganda, and Takataka Plastics. For the work carried out in IIT Bombay, we sincerely thank and recognize the in-kind support from IIT Bombay CTARA (Prof. Bakul Rao, Sushma Kulkarni, Dr. Chandana N, and Hemlata Suryawanshi).

We are thankful to the UC Berkeley students of Spring 2021 and 2022 cohorts who participated in the Sustainable Design for Developing Communities course and facilitated the development of user-friendly instruction guides for open access availability on the Gadgil Lab’s website. We are thankful to Michael Gee for his assistance on laboratory experiments at UC Berkeley, and to Emilie Kathol-Voilleque for her input on the Electro-Clean project. We would also like to recognize and thank Amba Moses and Bridget Adoch from Takataka Plastics for operating and testing the Electro-Clean process in Uganda. Finally, we would like to thank Jun Yu who helped us with the design of the Electro-Clean logo through the Catchafire platform.

We would like to acknowledge the students Denisse Montes-Arias, Norma-Patricia Alvarez- Vargas, and Sonia-Fernanda Torres-Flores from the Higher Technological Institutes of Abasolo (ITESA) and Irapuato (ITESI) in Mexico for conducting the Electro-Clean process at a household scale and for creating Youtube tutorials on the process. Many thanks to the students at the Higher Technological Institute of Irapuato (ITESI) who participated in the Biochemistry class in the year 2020, taught by Professor Varinia Lopez-Ramirez, in further improving the Electro-Clean process. Finally, we would like to acknowledge the students David Emmanuel, Boma Bill Odor, and Godwin Adams at Covenant University in Nigeria for conducting laboratory experiments on the Electro-Clean process; and the technicians Mr. Stephen Ayegbo and Mr. Ayo Akindele for their support in making the Electro-Clean process possible at Covenant University.

## Competing interests

The authors declare that they have no known competing financial interests or personal relationships that could have appeared to influence the work reported in this paper.

## Supporting Information

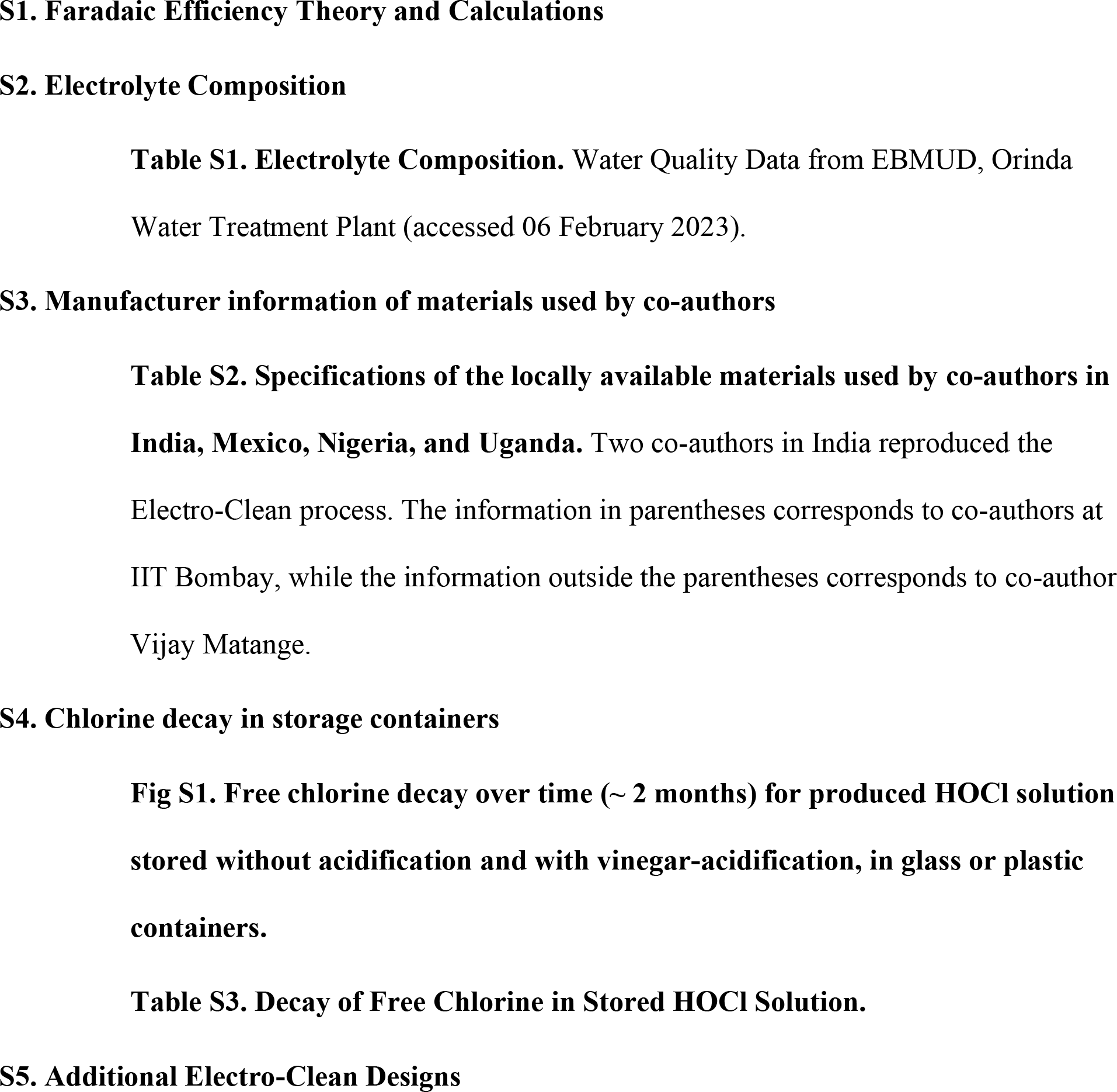
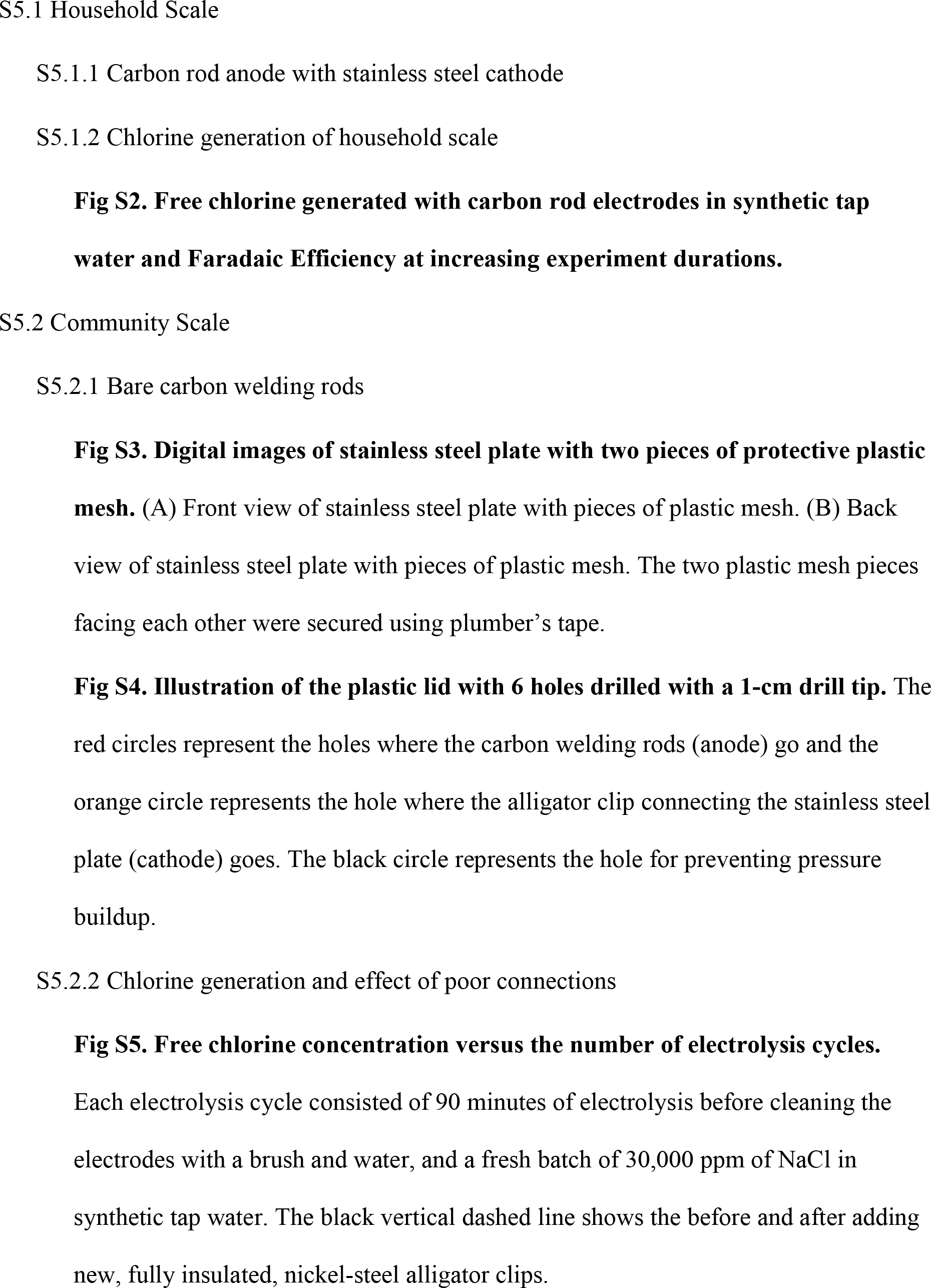
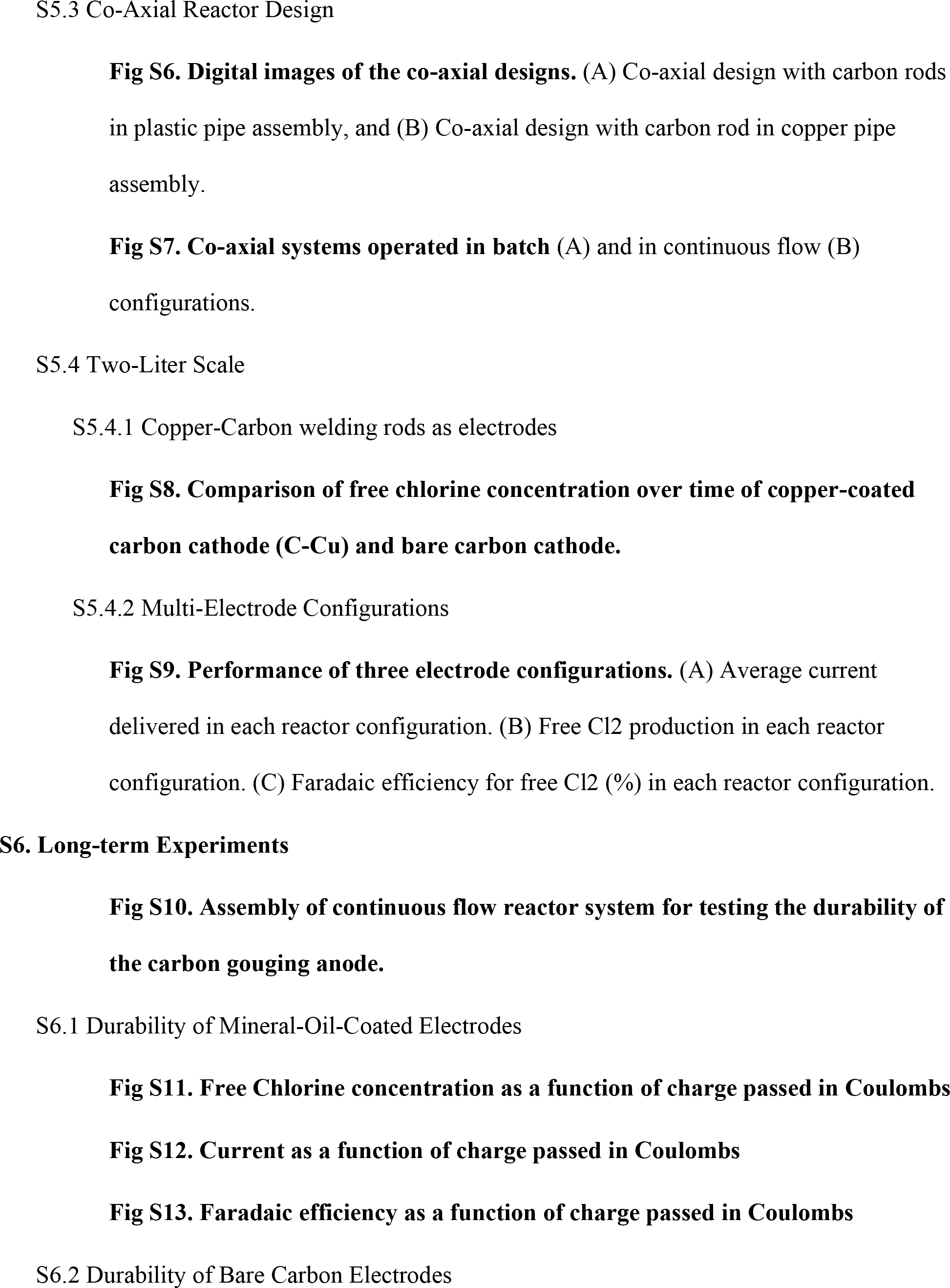
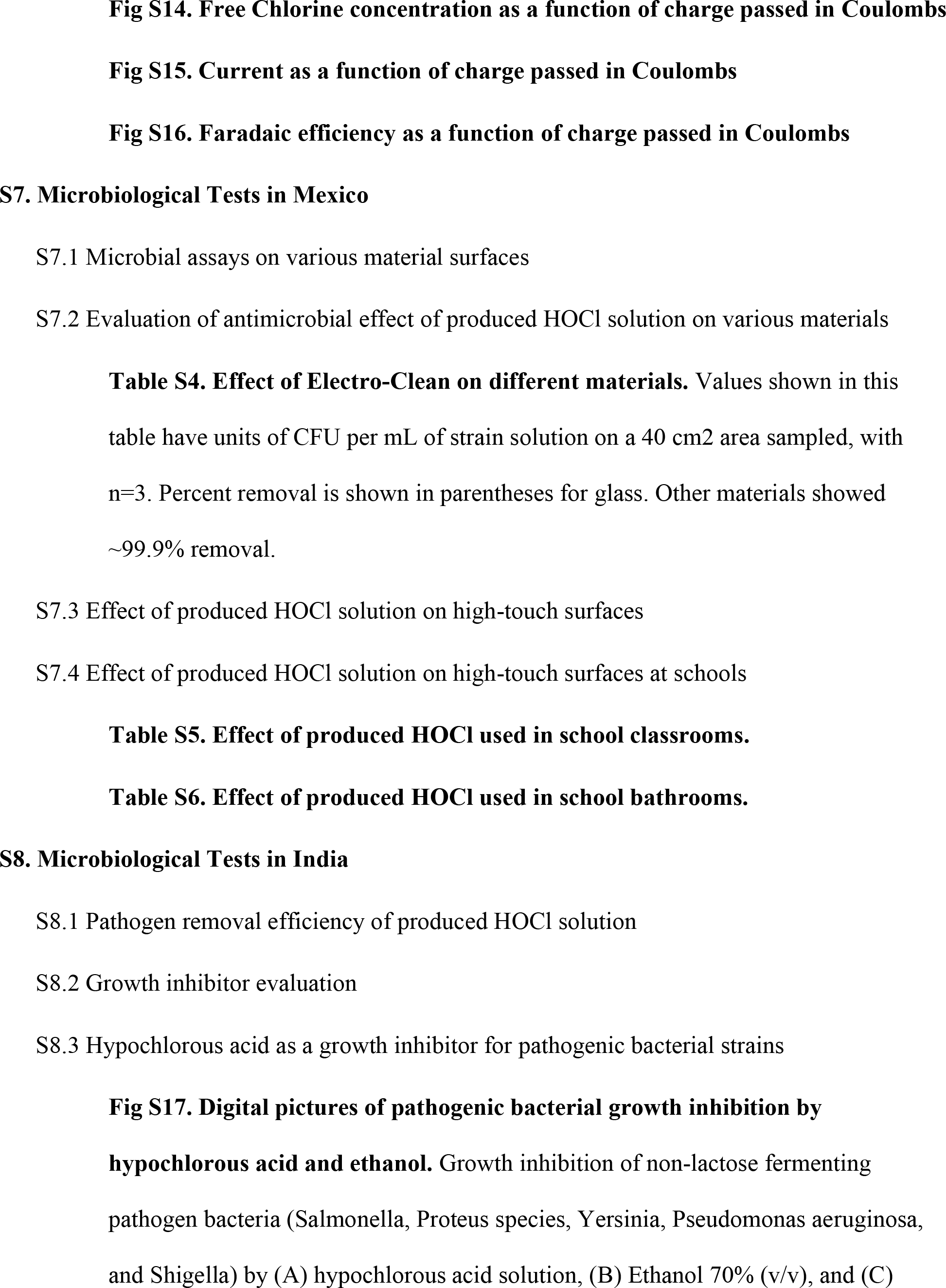
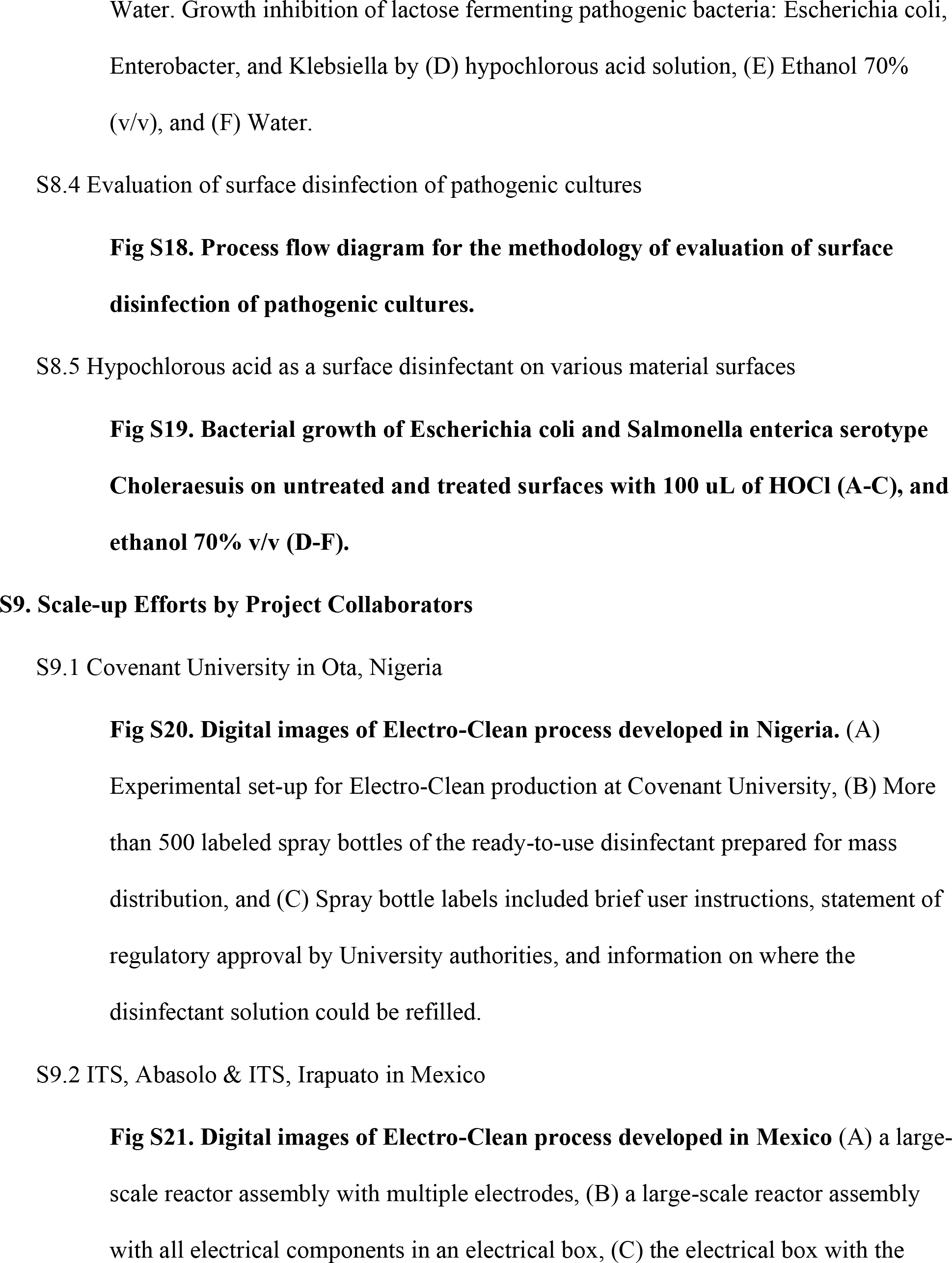
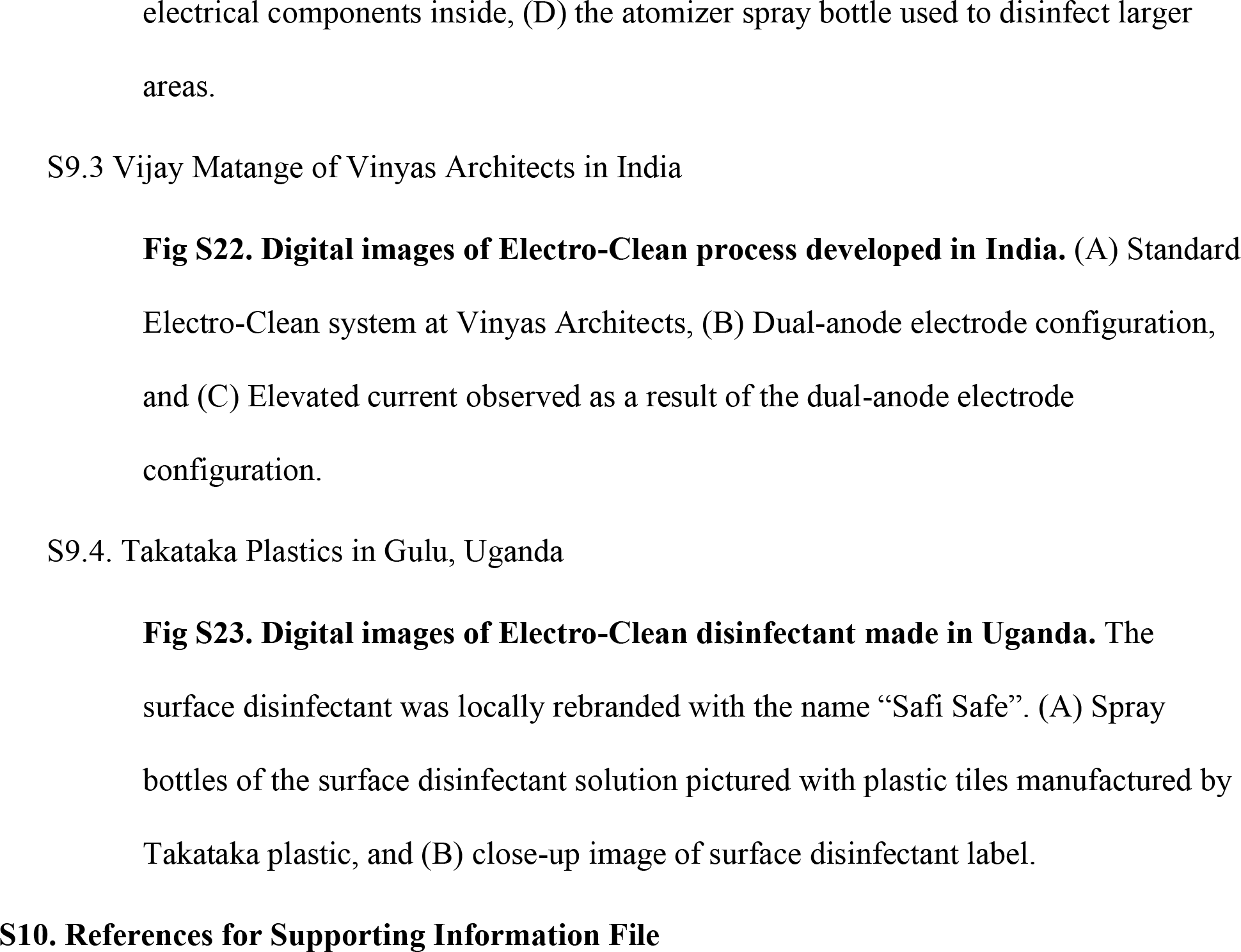

## Notes

### Competing Interest Statement

The authors have declared no competing interest.

### Funding Statement

There was partial financial support for this work from the Balaton Group, and Rudd Chair funds from Professor Ashok Gadgil. The Higher Technological Institutes of Abasolo and Irapuato provided additional financial support for the work described in Mexico. The NSF GRFP funded the graduate studies of authors Andrea Naranjo-Soledad and Dana Hernandez at UC Berkeley. The funders had no role in study design, data collection and analysis, decision to publish, or preparation of the manuscript.

### Author Declarations

Our study did not involve human subjects. No IRB was required.

